# Towards A Foundation Model for Clinical Voice Biomarkers

**DOI:** 10.64898/2026.05.28.26354346

**Authors:** Olivier Elemento, Alexandros Sigaras, Joseph T. Colonel, Iman Hajirasouliha, Satrajit S. Ghosh, Yael Bensoussan, Bridge2AI-Voice Consortium, Anaïs Rameau

**Author notes:** **Corresponding authors:** Olivier Elemento, Anaïs Rameau. A full list of Bridge2AI-Voice Consortium members and their affiliations appears at the end of the manuscript.

## Abstract

Vocal biomarkers, encompassing voice and speech [1], have largely been developed for individual conditions in isolation, limiting their generalizability across diseases and recording settings. To address this, we introduce VoiceFM, a contrastive model that learns general-purpose clinical voice representations by aligning audio embeddings with rich clinical metadata. Using the Bridge2AI-Voice dataset (984 primarily English-speaking adult participants — 846 used for training and 138 held out as a temporally separated validation cohort — 40,056 recordings totaling 176 hours across 5 academic medical centers), VoiceFM pairs a fine-tuned Whisper large-v2 encoder with a tabular transformer over 44 clinical features via symmetric InfoNCE loss. Linear probes on frozen VoiceFM embeddings achieve mean AUROC 0.952 ± 0.005 across five evaluation tasks (control vs disease screening plus four disease categories), significantly outperforming Frozen Whisper (0.926 ± 0.013, p = 0.013), Frozen HuBERT (0.885 ± 0.017, p = 0.0009), and the contrastively trained VoiceFM-HuBERT (0.938 ± 0.006, p = 0.012). On the 138-participant held-out cohort, VoiceFM-Whisper achieves AUROCs of 0.99 for Alzheimer’s/dementia/MCI and 0.89 for airway stenosis, demonstrating that the learned representations generalize to participants the model has never seen. VoiceFM representations transfer to three external datasets without retraining and improve few-shot classification. Recording task attribution identifies a small set of speech tasks that match or exceed the full battery’s performance, suggesting shorter screening protocols are feasible. Trained predominantly on English audio, VoiceFM transfers without fine-tuning to Spanish-language Parkinson’s disease (PD) detection (NeuroVoz, 107 participants, AUROC 0.93 ± 0.02), with the signal dominated by articulatory rather than phonatory features. A fine-tuned classifier achieves participant-level AUROC 0.87 (sustained 0.85, countdown 0.80) on the mPower smartphone study (585 held-out participants). Together, these results show that contrastive alignment between voice and rich clinical metadata can serve as the basis for a clinical voice foundation model, producing a single set of transferable representations that generalize across diseases, languages, recording conditions, and patients enrolled after model freeze.

## Introduction

Voice and speech, together with respiratory sounds, are among the most accessible digital biomarkers in medicine [1]. Acoustic data collection does not require specialized, expensive equipment or invasive intervention and instead can be captured remotely via instruments such as smartphones. Acoustic data encodes information about otolaryngologic, neuromuscular, respiratory, cognitive and inflammatory systems [2]. Accordingly, numerous reports have established associations between specific acoustic features and a variety of conditions, for example increased jitter and shimmer in Parkinson’s disease [3], altered speech rate and prosody in depression [4], and changes in vocal fold vibration patterns in laryngeal disorders [5]. These findings have motivated a growing number of studies on automated voice-based screening and monitoring, with promising results reported for conditions including Parkinson’s disease [6], Alzheimer’s disease [7], COVID-19 [8], and laryngeal pathology [9]. We have previously termed this growing field “audiomics” [10].

Despite this progress, the field remains fragmented, and the Food and Drug Administration (FDA) has yet to approve a voice biomarker for diagnostic purposes. Most voice diagnostic models are trained on small, single-condition datasets with narrow recording protocols, then evaluated on the same distribution [11]. This limits models’ predictive capabilities when multiple conditions are co-occurring and contributes to their limited generalizability on external datasets. Models built for Parkinson’s detection cannot screen for depression; those trained on sustained vowels do not generalize to connected speech. Beyond limited datasets and issues related to confounders, the fundamental limitation is architectural: each model encodes condition-specific features rather than a general representation of vocal health. This contrasts with the trajectory of other biomedical domains, where foundation models trained on large, diverse datasets have enabled multi-task transfer learning, from medical imaging [12] to electronic health records [13] to protein structure [14].

The emergence of self-supervised speech representations, particularly Hidden-Unit BERT (HuBERT) [15] and wav2vec 2.0 [16], has partially addressed the representation problem. These models learn general-purpose acoustic features from large unlabeled speech corpora and can be fine-tuned for downstream clinical tasks. However, they are trained on speech data from healthy speakers (primarily LibriSpeech), with limited exposure to pathological voice and no clinical context. More recently, Google’s Health Acoustic Representations (HeAR) model [17] has extended this paradigm to health-related sounds by pretraining a Vision Transformer on approximately 3.1 billion two-second audio clips extracted from YouTube videos using a health acoustic event detector. While HeAR increases acoustic diversity over speech-only corpora, it is still trained without clinical ground truth: the training data is weakly filtered for health-related sounds but carries no diagnosis, severity, or questionnaire information. The features that distinguish healthy from diseased speech may therefore remain underrepresented or absent in these generic representations.

To address this gap, we propose to use contrastive alignment between voice recordings and clinical metadata, including clinician-validated disease diagnoses (ground truth). Instead of training a model to classify specific conditions, we train it to align each recording with the full clinical profile of the speaker, including demographics, disease categories, validated questionnaire scores (Patient Health Questionnaire-9 [PHQ-9], Generalized Anxiety Disorder-7 [GAD-7], Voice Handicap Index-10 [VHI-10]), and clinician-rated perceptual assessments. The resulting model learns a voice representation that encodes acoustic features that predict the clinical state, without being told which conditions or features matter.

This approach is enabled by the Bridge2AI-Voice as a Biomarker of Health (B2AI-Voice) dataset [18], a multi-site clinical voice collection with unprecedented metadata richness. B2AI-Voice has enrolled close to 1,000 adult participants across five academic medical centers in the United States and Canada, following a 22-task standardized recording protocol per participant (sustained vowels, reading passages, spontaneous speech, diadochokinesis, cognitive-linguistic tasks; not all participants completed every task) alongside detailed clinical phenotyping. We trained VoiceFM on the 846 participants available at the time of model development, all of whom met a stringent gold-standard diagnosis (GSD) labeling protocol, and then evaluated it on a temporally separated cohort of 138 additional participants enrolled at the same five sites under the same protocol after training was finalized. Because this held-out cohort shares the recording infrastructure and protocol of the training cohort, it tests post-freeze generalization to new patients rather than distribution shift across sites or instruments — that test is provided separately by the external datasets and the NeuroVoz / mPower analyses below. The combination of diverse recording protocols, rich clinical metadata, and a separately enrolled validation cohort makes B2AI-Voice uniquely suited for training and validating a voice foundation model.

We present VoiceFM, a Contrastive Language-Image Pretraining (CLIP)-style [19] dual-encoder model that pairs a fine-tuned speech encoder with a tabular transformer [20] over clinical features, trained with symmetric Noise-Contrastive Estimation (InfoNCE) loss [21] and auxiliary objectives. We evaluate two audio backbones (Whisper large-v2 [22] and HuBERT-base [15]) and assess VoiceFM on multi-condition classification, few-shot transfer to external datasets, cross-site generalization, recording task attribution, cross-lingual Parkinson’s disease (PD) detection (NeuroVoz, 107 Spanish-speaking participants), large-scale PD detection (mPower, 5,044 participants), and embedding interpretability.

## Results

### Cohort and recording characteristics

After applying the GSD labeling protocol to the most recent B2AI-Voice data release (Methods), we retained 984 participants enrolled across five academic medical centers in the United States and Canada. VoiceFM was trained on the 846 participants available at the time of model development (**Table 1**), used for all model fitting and seed-based cross-validation; 138 additional participants were enrolled into B2AI-Voice after training was finalized and held out as an independent validation cohort for an out-of-sample generalization test (**Table 2**). Across the full cohort, participants span four disease categories derived from clinician-validated gold-standard diagnosis (GSD) flags — voice disorders (n = 333), neurodegenerative conditions (n = 227), respiratory disease (n = 191), mood disorders (n = 95) — plus a control group (n = 199); a category is positive when any of its constituent GSDs is positive (e.g., cat_neuro = 1 iff GSD-Parkinson’s, Alzheimer’s/MCI, Huntington’s, or ALS). 60 participants carry GSD diagnoses across two or more disease categories, so category counts sum to more than the cohort size. The cohort is 61% female, with a mean age of 59.6 years (SD 18.3). Validated clinical instruments include the Patient Health Questionnaire (PHQ-9, mean 4.2), Generalized Anxiety Disorder scale (GAD-7, mean 3.6), and Voice Handicap Index (VHI-10, mean 11.4).

**Table 1.** Training cohort characteristics by disease category. Demographic and clinical characteristics of the training cohort (N = 846), stratified by disease category and used to fit the contrastive VoiceFM model. Continuous variables (age, PHQ-9, GAD-7, VHI-10) reported as mean ± standard deviation; categorical variables as N (%). Disease categories are not mutually exclusive at the participant level. Disease assignment uses clinically-curated gold-standard diagnosis (GSD) criteria.

| Characteristic | Control | Voice | Neuro | Mood | Respiratory | Total |
| --- | --- | --- | --- | --- | --- | --- |
| N | 161 | 287 | 204 | 83 | 160 | 846 |
| Age, mean ± SD | 45.1 ± 20.0 | 62.1 ± 14.6 | 73.3 ± 9.6 | 49.5 ± 24.8 | 59.2 ± 13.1 | 59.4 ± 18.4 |
| Female | 89 (55.3%) | 188 (65.5%) | 91 (44.6%) | 48 (57.8%) | 126 (78.8%) | 514 (60.8%) |
| Male | 72 (44.7%) | 99 (34.5%) | 113 (55.4%) | 30 (36.1%) | 34 (21.2%) | 327 (38.7%) |
| Non-binary | 0 (0.0%) | 0 (0.0%) | 0 (0.0%) | 5 (6.0%) | 0 (0.0%) | 5 (0.6%) |
| Other | 0 (0.0%) | 0 (0.0%) | 0 (0.0%) | 0 (0.0%) | 0 (0.0%) | 0 (0.0%) |
| Hispanic/Latino | 28 (17.4%) | 24 (8.4%) | 3 (1.5%) | 6 (7.2%) | 9 (5.6%) | 69 (8.2%) |
| White | 94 (58.4%) | 235 (81.9%) | 169 (82.8%) | 67 (80.7%) | 141 (88.1%) | 661 (78.1%) |
| Black/African American | 23 (14.3%) | 28 (9.8%) | 8 (3.9%) | 3 (3.6%) | 10 (6.2%) | 71 (8.4%) |
| Asian | 23 (14.3%) | 12 (4.2%) | 11 (5.4%) | 10 (12.0%) | 3 (1.9%) | 58 (6.9%) |
| Indigenous | 4 (2.5%) | 5 (1.7%) | 1 (0.5%) | 1 (1.2%) | 2 (1.2%) | 12 (1.4%) |
| Pacific Islander | 1 (0.6%) | 2 (0.7%) | 1 (0.5%) | 0 (0.0%) | 0 (0.0%) | 4 (0.5%) |
| Other race | 22 (13.7%) | 14 (4.9%) | 17 (8.3%) | 5 (6.0%) | 8 (5.0%) | 63 (7.4%) |
| No formal/some HS | 4 (2.5%) | 7 (2.4%) | 19 (9.3%) | 7 (8.4%) | 2 (1.2%) | 32 (3.8%) |
| HS–Associates | 41 (25.5%) | 123 (42.9%) | 52 (25.5%) | 23 (27.7%) | 78 (48.8%) | 304 (35.9%) |
| College | 66 (41.0%) | 82 (28.6%) | 80 (39.2%) | 31 (37.3%) | 45 (28.1%) | 289 (34.2%) |
| Graduate+ | 50 (31.1%) | 75 (26.1%) | 53 (26.0%) | 22 (26.5%) | 35 (21.9%) | 221 (26.1%) |
| Hearing difficulty | 3 (1.9%) | 28 (9.8%) | 34 (16.7%) | 9 (10.8%) | 6 (3.8%) | 70 (8.3%) |
| Cognition difficulty | 7 (4.3%) | 27 (9.4%) | 68 (33.3%) | 31 (37.3%) | 11 (6.9%) | 122 (14.4%) |
| Mobility difficulty | 12 (7.5%) | 48 (16.7%) | 114 (55.9%) | 28 (33.7%) | 32 (20.0%) | 204 (24.1%) |
| Self-care difficulty | 0 (0.0%) | 11 (3.8%) | 73 (35.8%) | 17 (20.5%) | 3 (1.9%) | 86 (10.2%) |
| Independent living difficulty | 2 (1.2%) | 24 (8.4%) | 90 (44.1%) | 29 (34.9%) | 11 (6.9%) | 127 (15.0%) |
| Ever smoked | 11 (6.8%) | 60 (20.9%) | 48 (23.5%) | 11 (13.3%) | 20 (12.5%) | 139 (16.4%) |
| PHQ-9, mean ± SD | 2.2 ± 2.8 | 3.7 ± 4.1 | 6.2 ± 4.8 | 9.7 ± 5.8 | 3.4 ± 4.3 | 4.3 ± 4.7 |
| GAD-7, mean ± SD | 2.3 ± 3.2 | 3.3 ± 4.4 | 4.6 ± 4.8 | 8.4 ± 5.4 | 2.9 ± 4.1 | 3.7 ± 4.6 |
| VHI-10, mean ± SD | 2.7 ± 4.2 | 18.7 ± 8.9 | 9.9 ± 8.9 | 7.2 ± 7.9 | 11.3 ± 9.2 | 11.4 ± 10.1 |
| USF | 81 (50.3%) | 214 (74.6%) | 41 (20.1%) | 1 (1.2%) | 94 (58.8%) | 426 (50.4%) |
| Mt. Sinai | 0 (0.0%) | 0 (0.0%) | 160 (78.4%) | 34 (41.0%) | 3 (1.9%) | 160 (18.9%) |
| VUMC | 16 (9.9%) | 73 (25.4%) | 3 (1.5%) | 5 (6.0%) | 62 (38.8%) | 152 (18.0%) |
| WCM | 62 (38.5%) | 0 (0.0%) | 0 (0.0%) | 1 (1.2%) | 1 (0.6%) | 64 (7.6%) |
| MIT | 2 (1.2%) | 0 (0.0%) | 0 (0.0%) | 42 (50.6%) | 0 (0.0%) | 44 (5.2%) |
| Recordings, N | 6021 | 10826 | 11168 | 3515 | 5322 | 34232 |

**Table 2.** Validation cohort characteristics. Demographic and clinical characteristics of the validation cohort (N = 138), enrolled into B2AI Voice after the training cohort was finalized and evaluated only at inference time (see Figure 3). Same labeling criteria and reporting conventions as Table 1.

| Characteristic | Control | Voice | Neuro | Mood | Respiratory | Total |
| --- | --- | --- | --- | --- | --- | --- |
| N | 38 | 46 | 23 | 12 | 31 | 138 |
| Age, mean ± SD | 54.6 ± 18.8 | 60.7 ± 17.2 | 75.7 ± 7.0 | 67.9 ± 20.2 | 60.9 ± 16.4 | 60.9 ± 17.9 |
| Female | 21 (55.3%) | 31 (67.4%) | 11 (47.8%) | 7 (58.3%) | 18 (58.1%) | 82 (59.4%) |
| Male | 17 (44.7%) | 15 (32.6%) | 12 (52.2%) | 5 (41.7%) | 13 (41.9%) | 56 (40.6%) |
| Non-binary | 0 (0.0%) | 0 (0.0%) | 0 (0.0%) | 0 (0.0%) | 0 (0.0%) | 0 (0.0%) |
| Other | 0 (0.0%) | 0 (0.0%) | 0 (0.0%) | 0 (0.0%) | 0 (0.0%) | 0 (0.0%) |
| Hispanic/Latino | 5 (13.2%) | 10 (21.7%) | 1 (4.3%) | 0 (0.0%) | 6 (19.4%) | 21 (15.2%) |
| White | 27 (71.1%) | 38 (82.6%) | 16 (69.6%) | 9 (75.0%) | 29 (93.5%) | 110 (79.7%) |
| Black/African American | 3 (7.9%) | 5 (10.9%) | 2 (8.7%) | 1 (8.3%) | 1 (3.2%) | 11 (8.0%) |
| Asian | 4 (10.5%) | 0 (0.0%) | 1 (4.3%) | 0 (0.0%) | 0 (0.0%) | 5 (3.6%) |
| Indigenous | 1 (2.6%) | 1 (2.2%) | 1 (4.3%) | 1 (8.3%) | 0 (0.0%) | 3 (2.2%) |
| Pacific Islander | 0 (0.0%) | 1 (2.2%) | 0 (0.0%) | 0 (0.0%) | 0 (0.0%) | 1 (0.7%) |
| Other race | 4 (10.5%) | 3 (6.5%) | 4 (17.4%) | 2 (16.7%) | 1 (3.2%) | 12 (8.7%) |
| No formal/some HS | 0 (0.0%) | 1 (2.2%) | 3 (13.0%) | 2 (16.7%) | 1 (3.2%) | 5 (3.6%) |
| HS—Associates | 14 (36.8%) | 21 (45.7%) | 4 (17.4%) | 4 (33.3%) | 11 (35.5%) | 50 (36.2%) |
| College | 12 (31.6%) | 14 (30.4%) | 11 (47.8%) | 2 (16.7%) | 5 (16.1%) | 42 (30.4%) |
| Graduate+ | 12 (31.6%) | 10 (21.7%) | 5 (21.7%) | 4 (33.3%) | 14 (45.2%) | 41 (29.7%) |
| Hearing difficulty | 4 (10.5%) | 4 (8.7%) | 7 (30.4%) | 4 (33.3%) | 3 (9.7%) | 18 (13.0%) |
| Cognition difficulty | 0 (0.0%) | 4 (8.7%) | 10 (43.5%) | 5 (41.7%) | 0 (0.0%) | 15 (10.9%) |
| Mobility difficulty | 0 (0.0%) | 11 (23.9%) | 15 (65.2%) | 9 (75.0%) | 7 (22.6%) | 32 (23.2%) |
| Self-care difficulty | 0 (0.0%) | 2 (4.3%) | 6 (26.1%) | 3 (25.0%) | 2 (6.5%) | 10 (7.2%) |
| Independent living difficulty | 0 (0.0%) | 3 (6.5%) | 6 (26.1%) | 3 (25.0%) | 1 (3.2%) | 11 (8.0%) |
| Ever smoked | 0 (0.0%) | 0 (0.0%) | 0 (0.0%) | 0 (0.0%) | 0 (0.0%) | 0 (0.0%) |
| PHQ-9, mean ± SD | 2.1 ± 2.2 | 3.8 ± 5.0 | 5.1 ± 3.0 | 7.2 ± 3.9 | 4.3 ± 5.7 | 3.6 ± 4.3 |
| GAD-7, mean ± SD | 2.0 ± 2.7 | 3.2 ± 4.4 | 4.1 ± 5.3 | 7.3 ± 7.1 | 3.5 ± 4.3 | 3.1 ± 4.3 |
| VHI-10, mean ± SD | 4.4 ± 5.9 | 19.6 ± 9.5 | 7.9 ± 8.4 | 9.3 ± 9.2 | 10.2 ± 8.0 | 11.1 ± 10.1 |
| USF | 24 (63.2%) | 33 (71.7%) | 2 (8.7%) | 0 (0.0%) | 27 (87.1%) | 83 (60.1%) |
| Mt. Sinai | 0 (0.0%) | 0 (0.0%) | 21 (91.3%) | 9 (75.0%) | 0 (0.0%) | 21 (15.2%) |
| VUMC | 0 (0.0%) | 13 (28.3%) | 0 (0.0%) | 1 (8.3%) | 4 (12.9%) | 18 (13.0%) |
| WCM | 12 (31.6%) | 0 (0.0%) | 0 (0.0%) | 0 (0.0%) | 0 (0.0%) | 12 (8.7%) |
| MIT | 2 (5.3%) | 0 (0.0%) | 0 (0.0%) | 2 (16.7%) | 0 (0.0%) | 4 (2.9%) |
| Recordings, N | 1453 | 2105 | 1013 | 493 | 1278 | 5824 |

Each participant followed a 22-task standardized recording protocol (some tasks include multiple prompt variants in the data, with not all participants completing every task). This yielded 40,056 recordings totaling 176 hours of audio across the cohort (**Table 3**). The 22 protocol tasks are grouped into 13 task categories that span six functional domains: sustained phonation (prolonged vowels, maximum phonation time), connected speech (reading passages, spontaneous narratives), diadochokinesis (DDK; rapid syllable repetition), cognitive-linguistic tasks (confrontation naming, verbal fluency, productive vocabulary), respiratory maneuvers (cough, breathing), and quality-control prompts. This diversity is essential for a foundation model, as different conditions likely manifest in different vocal behaviors, with no single recording type capturing all clinically relevant information.

**Table 3.** Recording battery composition. Composition of the recording battery (N = 40,056 recordings; 984 participants) grouped into 13 task categories. Recordings: number of recordings per category. Participants: number of participants contributing at least one recording in that category. Types: number of distinct recording prompts in the category. Mean Duration: average length per recording in seconds. Total Duration: cumulative time in hours.

| Task Category | Recordings | Participants | Types | Mean Duration (s) | Total Duration (h) |
| --- | --- | --- | --- | --- | --- |
| Free Speech / Narrative | 5978 | 975 | 20 | 37.6 | 62.4 |
| Respiration / Cough | 9449 | 970 | 27 | 15.0 | 39.5 |
| Sustained Phonation | 5764 | 979 | 15 | 11.9 | 19.1 |
| Diadochokinesis | 5275 | 974 | 10 | 6.1 | 8.9 |
| Harvard Sentences | 6100 | 296 | 720 | 4.1 | 6.9 |
| CAPE-V Sentences | 2562 | 425 | 18 | 3.7 | 2.6 |
| Productive Vocabulary | 1908 | 242 | 6 | 10.3 | 5.5 |
| Reading Passages | 1176 | 774 | 3 | 49.1 | 16.0 |
| Loudness Tasks | 1058 | 977 | 2 | 5.4 | 1.6 |
| Confrontation Naming | 311 | 237 | 1 | 75.6 | 6.5 |
| Verbal Fluency | 159 | 146 | 2 | 55.3 | 2.4 |
| Random Item Generation | 307 | 236 | 2 | 57.1 | 4.9 |
| Other | 9 | 9 | 1 | 9.8 | 0.0 |
| **Total** | **40056** | **984** | **827** | **15.8** | **176.3** |

### VoiceFM architecture and training

To implement VoiceFM’s CLIP-style dual-encoder architecture, we tested two audio backbones. We assessed Whisper large-v2 as encoder [22] (32 transformer layers, 1280 hidden dimensions, ∼637M parameters), with layers 0–27 frozen and layers 28–31 fine-tuned, followed by mean pooling and projection to 256 dimensions (∼643M total parameters, ∼85M trainable) (VoiceFM-Whisper, **Figure 1a**). We also evaluated HuBERT-base (12 transformer layers, 768 dimensions, 95M parameters), with layers 0–8 frozen and layers 9–11 fine-tuned, followed by attentive pooling and projection to 256 dimensions (∼99.2M total parameters, ∼37.9M trainable) (VoiceFM-HuBERT). Both models share the same clinical encoder, implemented as a tabular transformer [20] (4 layers, 4 attention heads) over 44 features (demographics, disease categories, validated questionnaire scores, functional status; **Table S1**) projected to 256 dimensions. We trained both models with the same contrastive objective, using symmetric InfoNCE loss with auxiliary disease-category and age-regression heads. We adopted hard-negative mining for the final models (see Methods, **Figure 1b,c** shows a representative training seed for VoiceFM-Whisper).

**Figure 1.**
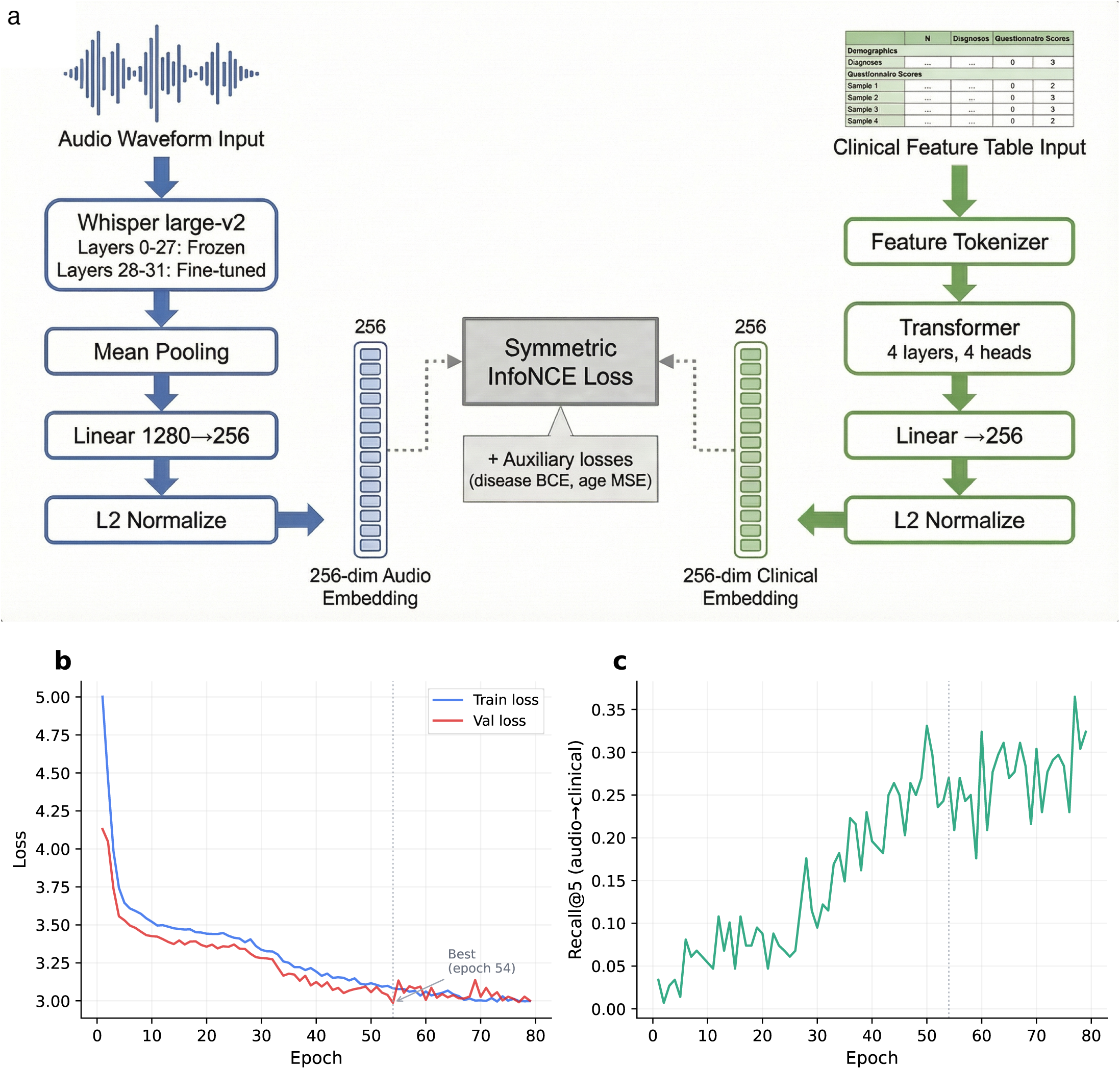
VoiceFM architecture and training. a, VoiceFM dual-encoder architecture: a fine-tuned Whisper large-v2 audio encoder (layers 28–31 unfrozen) produces 256-dimensional audio embeddings aligned with clinical embeddings from a tabular transformer over 44 clinical features via symmetric InfoNCE loss, with auxiliary disease-category BCE and age regression MSE losses. b, Training loss (train and validation) across epochs for VoiceFM-Whisper (representative seed). c, Retrieval performance (Recall@5, audio-to-clinical) across epochs. Trained on 846 participants; early stopping with patience 25.

### Disease classification from voice embeddings

We evaluated VoiceFM by training linear probes (logistic regression) on frozen 256-dimensional audio embeddings using gold-standard diagnosis labels, comparing VoiceFM-Whisper and VoiceFM-HuBERT against their respective frozen baselines (**Figure 2a**). All four models were evaluated under identical methodology, and all results are 5-seed means with stratified participant-level train/test splits (70/15/15).

**Figure 2.**
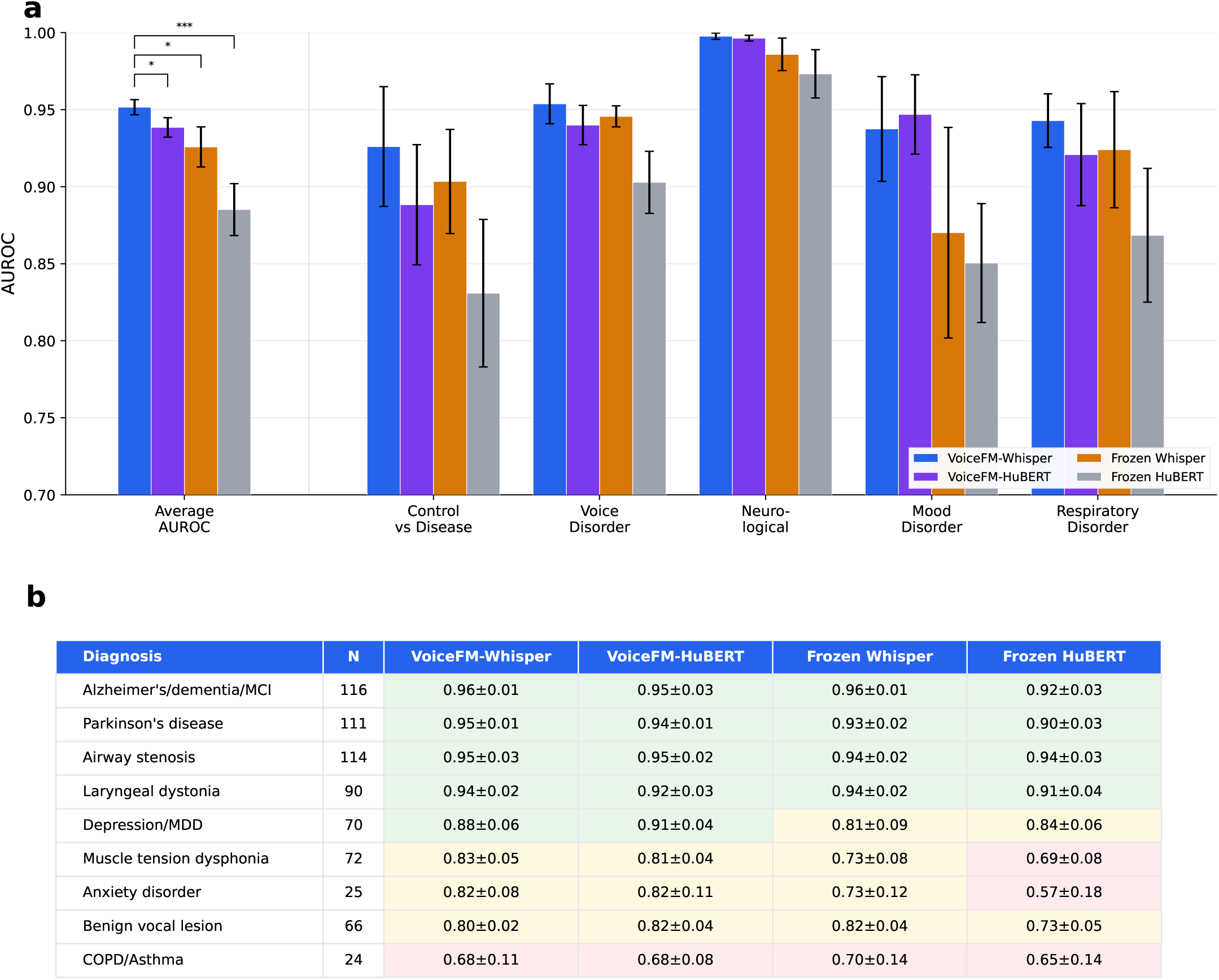
GSD classification performance. a, Mean AUROC across five gold-standard diagnosis (GSD) categories for four models — VoiceFM-Whisper, VoiceFM-HuBERT, Frozen Whisper, Frozen HuBERT. Left group shows the average across categories; right groups show per-category performance. Error bars: SD across 5 seeds (42–46). Significance markers: Welch’s t-test on the seed-level AUROC distributions. b, Per-diagnosis AUROC for 9 individual GSD conditions (test-fold positives ≥ 10 per seed).

On the five evaluation tasks (four disease categories, plus control vs disease screening), VoiceFM-Whisper achieved mean AUROC 0.952 ± 0.005. It outperformed Frozen Whisper (0.926 ± 0.013, Welch’s t-test p = 0.013), VoiceFM-HuBERT (0.938 ± 0.006, p = 0.012) and Frozen HuBERT (0.885 ± 0.017, p = 0.0009). VoiceFM-HuBERT also significantly outperformed Frozen HuBERT (p = 0.0018). VoiceFM-Whisper achieved strongest per-category performance on neurological conditions (0.998 ± 0.002), followed by voice disorders (0.954 ± 0.013), respiratory disease (0.943 ± 0.017), mood disorders (0.937 ± 0.034), and control versus disease (0.926 ± 0.039).

In a per-diagnosis analysis (**Figure 2b**), VoiceFM-Whisper achieved strong performance for Alzheimer’s/MCI (0.961 ± 0.007), Parkinson’s disease (0.951 ± 0.011), airway stenosis (0.950 ± 0.033), laryngeal dystonia (0.943 ± 0.021), and depression (0.880 ± 0.055). Performance was weakest for COPD/asthma (0.682 ± 0.112), a heterogeneous category that combines two distinct respiratory conditions.

### Backbone and training strategy comparison

The four-model comparison in **Figure 2a** and **2b** revealed two things. First, contrastive fine-tuning significantly improves both backbones: VoiceFM-Whisper outperformed Frozen Whisper (p = 0.013) and VoiceFM-HuBERT outperformed Frozen HuBERT (p = 0.0018). Second, the backbone choice matters independently: Frozen Whisper already significantly outperformed Frozen HuBERT (p = 0.006), and VoiceFM-HuBERT did not significantly exceed Frozen Whisper (p = 0.13). This indicates that Whisper’s larger pre-trained representation (1280 dimensions, 32 layers, trained on 680,000 hours of multilingual audio) provides a stronger starting point than HuBERT (768 dimensions, 12 layers, trained on LibriSpeech). Importantly, our results demonstrate that contrastive fine-tuning improves over the strong foundations provided by both audio models. We also assessed VoiceFM-HeAR (AUROC 0.922), which uses a frozen Google HeAR encoder [17] with only the projection layer trainable, and a Frozen HeAR baseline (AUROC 0.874) (**Figure S1**). Contrastive alignment improved all three backbones over their frozen baselines (Whisper +0.026, HuBERT +0.054, HeAR +0.049). As baseline control, we also performed a simple logistic regression on age and sex alone. This baseline approach achieved mean AUROC 0.65 across the five categories (highest in the neurological category at 0.83, where age is a known risk factor for both PD and AD/MCI), lower than all audio-based models (**Figure S1**).

### Evaluation on a temporally held-out cohort

The five-seed evaluation above (five independent runs, each using a fresh stratified 70/15/15 participant-level train/validation/test split) evaluated the model under standard in-distribution conditions. To assess out-of-sample generalization, we ran inference on the independent validation cohort of 138 participants (38 controls, 46 voice, 23 neurological, 12 mood, 31 respiratory; **Table 2**). These participants were enrolled into B2AI-Voice after the training cohort was finalized and were never seen during model fitting or seed-based cross-validation. Linear probes on both VoiceFM-Whisper and VoiceFM-HuBERT embeddings achieved strong AUROCs across disease categories (**Figure 3**). For VoiceFM-Whisper, we observed control vs disease 0.910 ± 0.005, voice 0.964 ± 0.023, neurological 0.984 ± 0.024, mood 0.849 ± 0.051, respiratory 0.832 ± 0.013 (**Figure 3a**). In a per-diagnosis analysis (**Figure 3b**), Alzheimer’s/dementia/MCI (0.986 ± 0.006) and Parkinson’s disease (0.928 ± 0.020) performed best, followed by airway stenosis (0.891 ± 0.024), laryngeal dystonia (0.890 ± 0.033), depression (0.841 ± 0.076), anxiety (0.818 ± 0.072), benign vocal lesion (0.809 ± 0.096), muscle tension dysphonia (0.772 ± 0.069), and COPD/asthma (0.699 ± 0.091). Good but overall lower AUROCs were observed for VoiceFM-HuBERT (**Figure 3a, 3b**). We note that this cohort shares the recording infrastructure and clinical protocol of the training cohort, so it tests temporal generalization rather than distribution shift.

**Figure 3.**
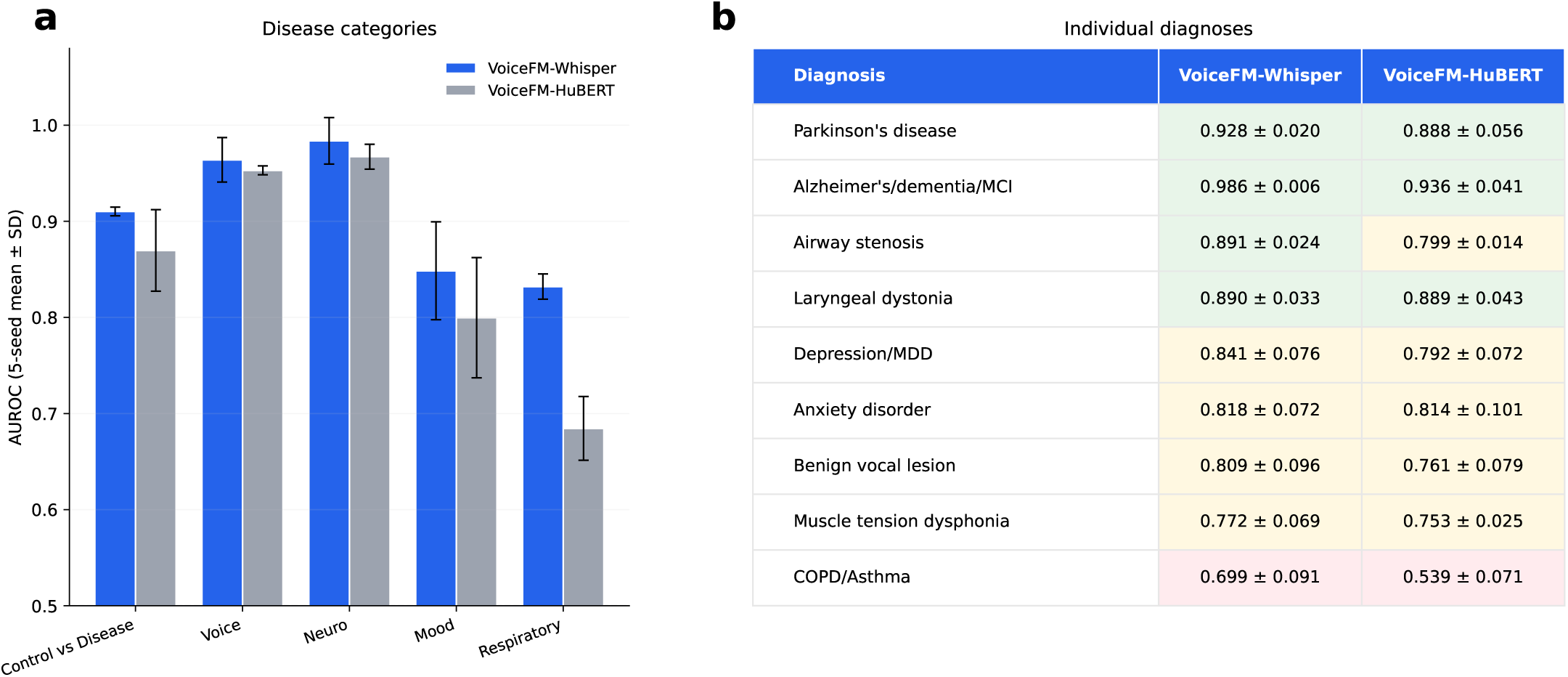
Prospective evaluation on held-out cohort. Out-of-sample evaluation on the 138 participants held out from training, enrolled into B2AI Voice after the training cohort was finalized. a, Disease-category AUROCs for VoiceFM-Whisper (blue) vs VoiceFM-HuBERT (gray); 5-seed mean ± SD (seeds 42–46). The held-out cohort is fixed across seeds; reported variance reflects probe sensitivity to the training-cohort split. b, Per-diagnosis AUROCs for 9 individual GSD conditions, sorted by VoiceFM-Whisper performance. Cell shading: green ≥ 0.85, yellow 0.70–0.85, red < 0.70.

### Transfer to external datasets

To test whether VoiceFM representations generalize beyond the B2AI-Voice cohort, we evaluated frozen VoiceFM-Whisper embeddings on three external datasets (**Figure 4a**): MDVR-KCL (Parkinson’s disease from voice recordings, n = 73), Saarbrücken Voice Database (SVD; voice pathology from sustained vowels, n = 2,041), and Coswara (COVID-19 from cough and breathing sounds, n = 2,098). Linear probes on frozen VoiceFM-Whisper embeddings outperformed Frozen Whisper on all three datasets: MDVR-KCL 0.905 ± 0.010 (vs 0.839 Frozen Whisper), SVD 0.788 ± 0.003 (vs 0.736), Coswara 0.764 ± 0.004 (vs 0.743).

**Figure 4.**
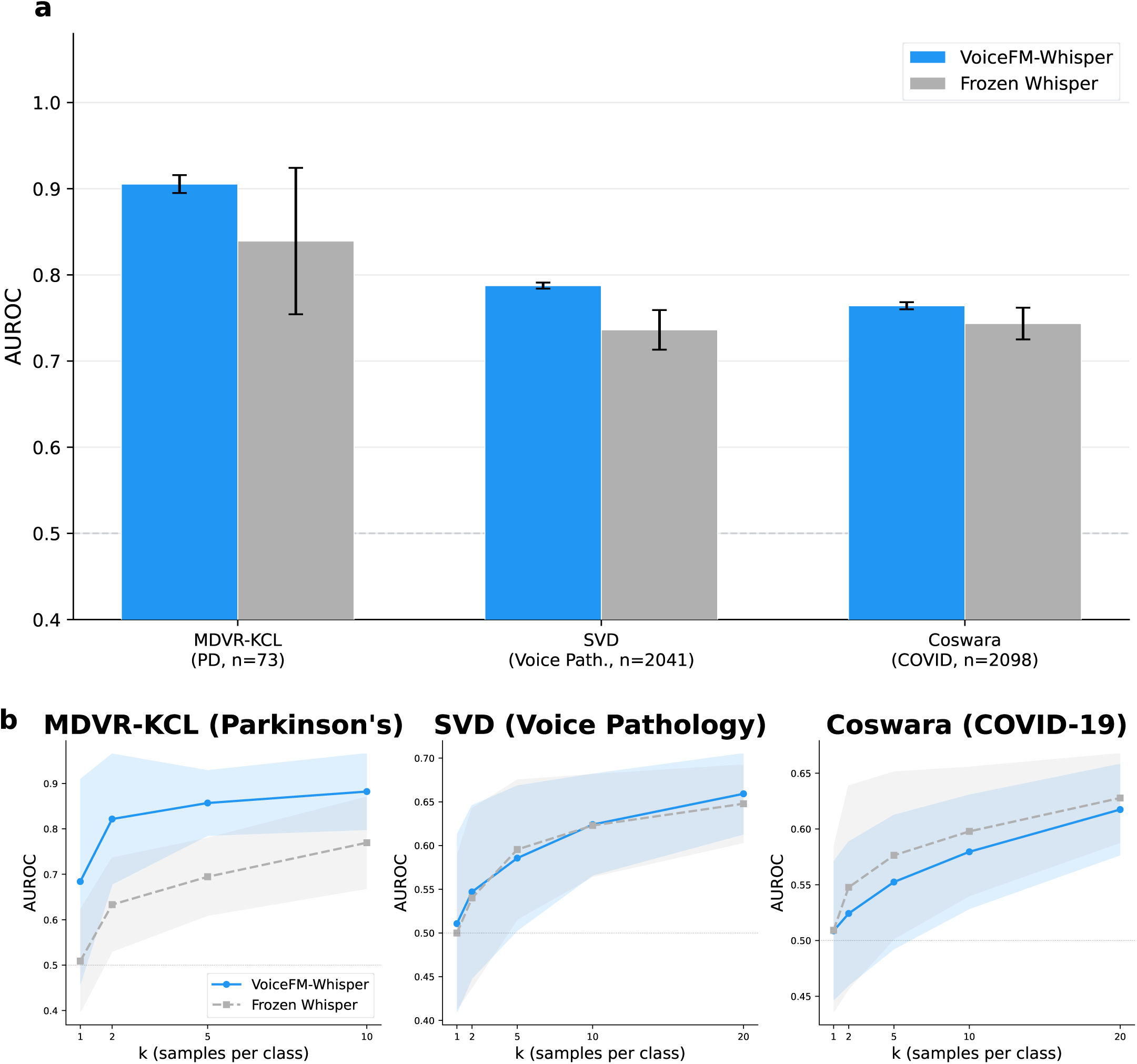
External dataset transfer and few-shot learning. a, Transfer AUROC on three external voice datasets not seen during training: Coswara (COVID-19 detection, n=2,098), Saarbrücken Voice Database (SVD; voice pathology, n=2,041), and MDVR-KCL (Parkinson’s disease, n=73). VoiceFM-Whisper (blue) vs Frozen Whisper (gray). 5-fold cross-validation with logistic regression probes on frozen embeddings. Error bars: SD across 5 seeds. b, Few-shot learning curves (k=1 to 20 labeled examples per class, 100 random trials per k). Shaded bands show standard deviation of the AUROC distribution: for VoiceFM-Whisper, pooled across (5 seeds × 100 trials) using the law of total variance (within-trial + between-seed); for Frozen Whisper (deterministic encoder), across the 100 trials. Both bands therefore measure sensitivity to support-set sampling and are directly comparable.

We also evaluated few-shot transfer performance across k ∈ {1, 2, 5, 10, 20} labeled examples per class (**Figure 4b**). For each k, we sampled k participants per class, then fitted a logistic-regression probe on their frozen 256-d embeddings, evaluated AUROC on the remaining participants. We performed this procedure across 100 random samplings and averaged the AUROCs. On MDVR-KCL, a Parkinson’s disease dataset, VoiceFM-Whisper showed a large advantage, reaching 0.82 AUROC with just k = 2 examples per class versus 0.63 for Frozen Whisper at the same k. On the other hand, on SVD and Coswara, the few-shot difference between VoiceFM-Whisper and Frozen Whisper was small and inconsistent across k. Altogether these results indicate that contrastive pre-training’s data-efficiency benefit is most pronounced on some tasks, e.g. the PD task. This is possibly explained by the high prevalence of neurological diagnoses in B2AI-Voice, and the baseline good performance of VoiceFM-Whisper in this disease class.

### Recording task attribution

We wondered whether all recording tasks contribute equally to disease classification or whether a few carry most of the signal. To address this, we first trained an independent logistic-regression probe for each recording type and ranked them by AUROC per disease category. This analysis showed that cognitive-linguistic tasks (story recall, verbal fluency, random item generation), connected reading, and spontaneous speech consistently ranked highest for neurological condition detection, while sustained phonation and respiratory tasks ranked lower for most disease categories (**Figure 5a**).

**Figure 5.**
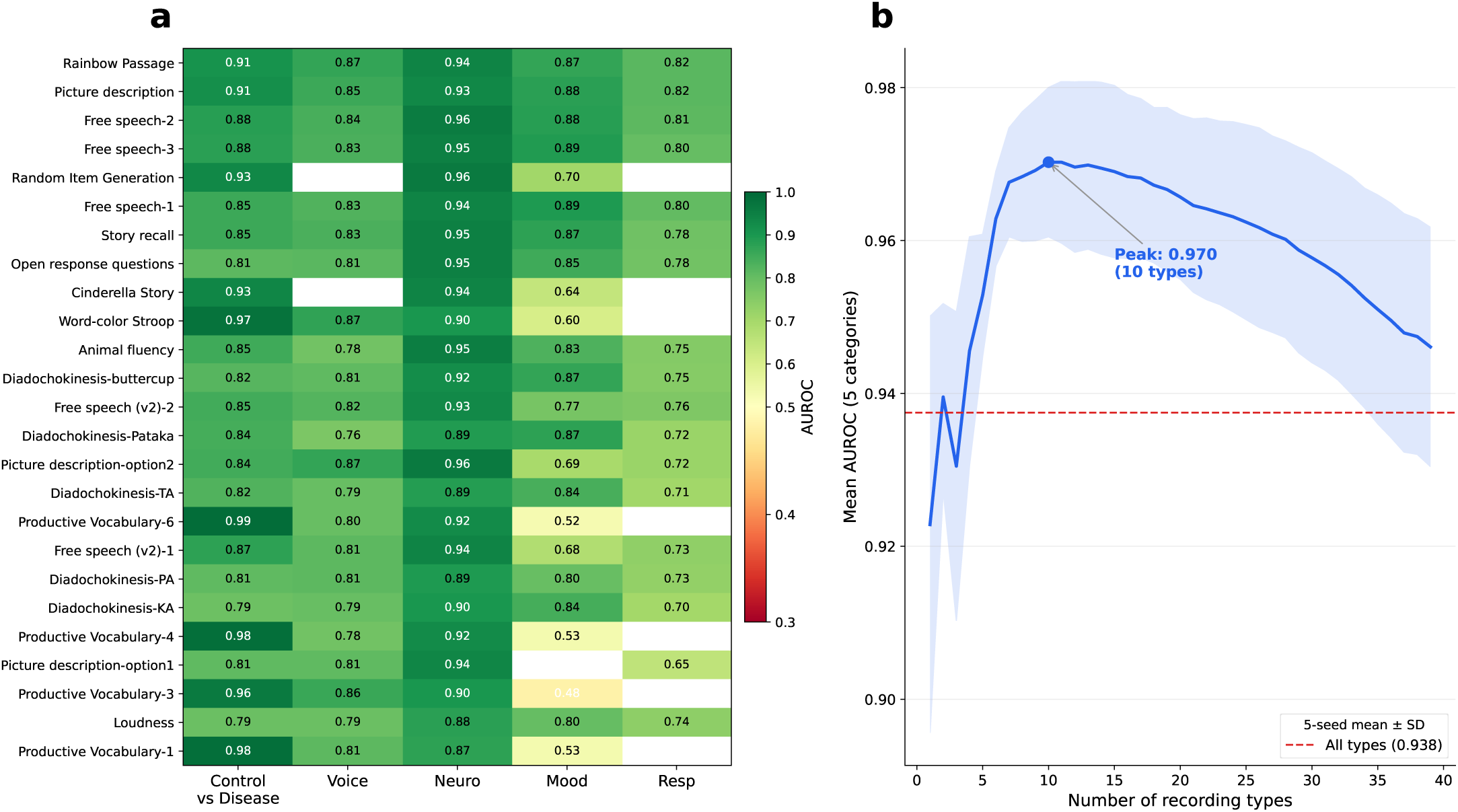
Recording task attribution. a, Per-recording-type AUROC heatmap for the top recording types across the five disease categories (Control vs Disease, Voice, Neurological, Mood, Respiratory). Blank cells indicate recording type × disease combinations where AUROC could not be reliably estimated due to insufficient test participants. b, Greedy forward selection on recording types: starting from no recordings, iteratively add the task type that most increases mean AUROC across the five categories. Solid line: 5-seed mean; shaded band: ± SD. The all-types baseline is shown as a red dashed line. Selection is performed on the same test set used for evaluation, so absolute peak AUROC may be optimistically biased; relative ranking of task types is the primary finding. A nested cross-validation sensitivity analysis (greedy selection signal taken from the validation split rather than the test split; data not shown) confirmed that the val-selected subset matches the all-types AUROC within seed variability, supporting the interpretation that a brief targeted protocol may match — but does not robustly exceed — the full battery.

We then ran greedy forward selection across recording types to find the minimal set that maximized mean classification AUROC across disease categories (**Figure 5b**; see Methods). We found that performance peaked at ∼0.97 AUROC across the five categories, slightly exceeding the all-types baseline of ∼0.94 (**Figure 5b**). On average, peak AUROC was achieved with only 10 recording types spanning complementary speech dimensions. This included free speech (which represents spontaneous language production), diadochokinesis (assessing rapid motor coordination), random item generation (executive function), and productive vocabulary (lexical retrieval). These results suggest that a targeted protocol covering a subset of the 22 task families (about 10 recording types in our analysis) may match or even improve classification performance compared to the more comprehensive battery.

### Embedding interpretability

To verify that VoiceFM embeddings encode meaningful acoustic properties, we extracted 14 traditional voice quality measures (fundamental frequency [F0], jitter, shimmer, harmonics-to-noise ratio [HNR], cepstral peak prominence smoothed [CPPS], formants) from prolonged vowel recordings and tested whether they can be decoded from the 256-dimensional embeddings via Ridge regression (**Figure S2a**).

VoiceFM-Whisper achieved positive R² for all 14 acoustic features (mean R² = 0.21), with the best performance for HNR (R² = 0.49), CPPS (R² = 0.36), shimmer (R² = 0.36), F0 mean (R² = 0.32), and jitter (R² = 0.25). In contrast, Frozen Whisper failed to linearly decode most features (mean R² = −0.60), achieving positive R² for only 2 of 14 features (HNR and CPPS). Notably, VoiceFM-Whisper achieved this with 256-dimensional embeddings, compared to 1280 for Frozen Whisper. The improvement therefore cannot be attributed to greater embedding dimensionality. Principal component analysis (PCA) revealed that the first principal component (43% of variance) organized the embedding space by voice quality, separating healthy-voice markers (CPPS, HNR; both positively correlated with PC1) from pathological-voice markers (jitter, shimmer; both negatively correlated) (**Figure S2b**).

We next examined the geometry of the embedding space using two complementary analyses: nearest-neighbor retrieval and within-participant cosine similarity. Nearest-neighbor retrieval (k = 5) placed VoiceFM-Whisper neighbors in the same disease category 69% of the time versus 63% for Frozen Whisper neighbors (Welch’s t-test p = 3.0 × 10⁻⁵, n = 846), well above the 24% chance baseline (**Figure S3a**). We also observed that VoiceFM-Whisper embeddings of different recordings from the same participant were substantially more similar (mean cosine similarity 0.94 ± 0.02 across participants) than embeddings from different participants (0.84 ± 0.03) (**Figure S3b**). Both speaker identity and disease structure were therefore present in the same embedding space.

### Parkinson’s disease detection

We have so far established that VoiceFM embeddings encode clinically meaningful structure on the training cohort. We then sought to test whether that structure transfers to cohorts the model was never trained on, or recorded in languages and acoustic conditions absent from the pre-training distribution. We focused on Parkinson’s disease, because it has a well-characterized vocal signature and independent datasets in multiple languages. The latter include a large-scale longitudinal smartphone cohort useful for real-world evaluation (**Figure 6**).

**Figure 6.**
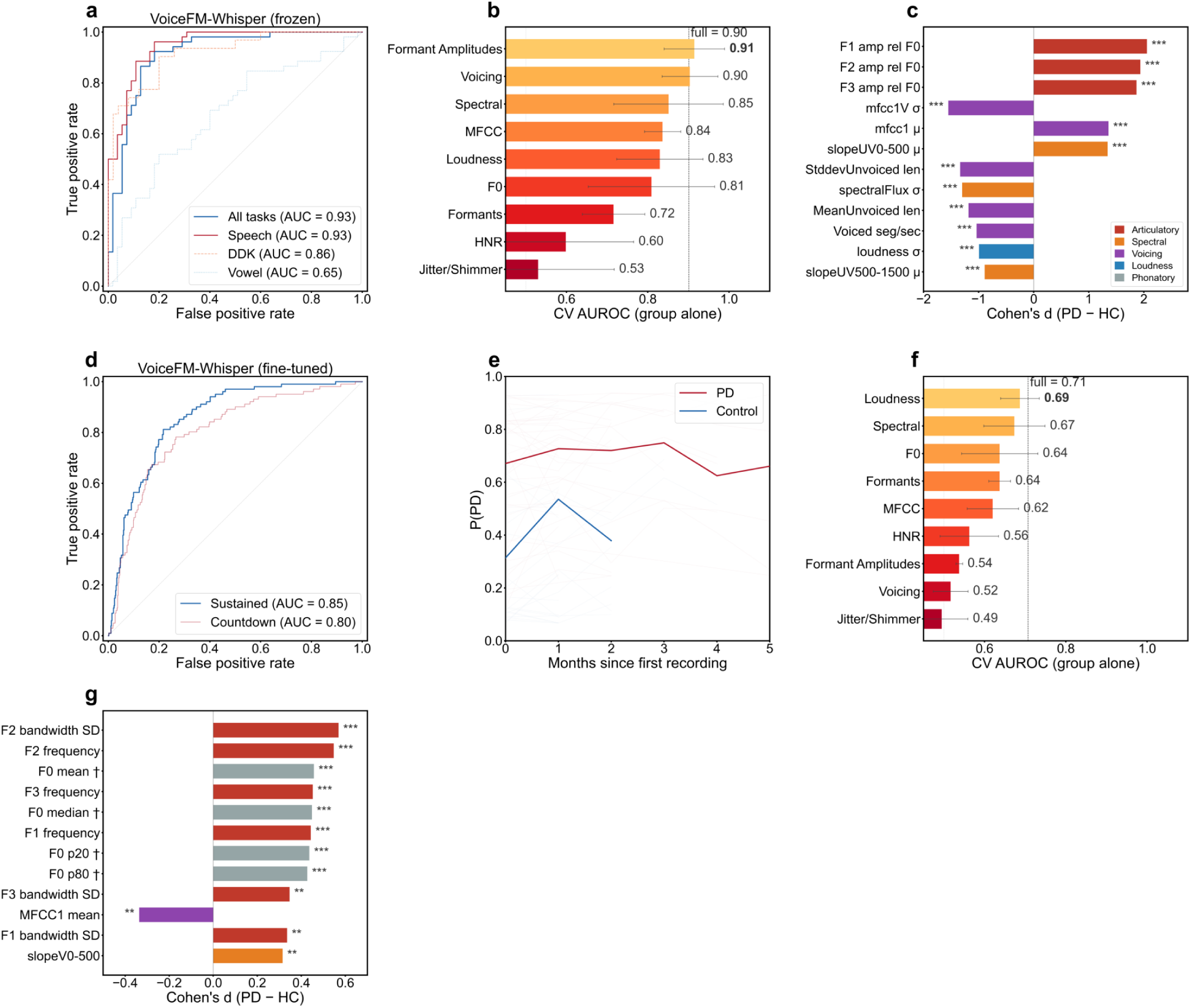
Application to Parkinson’s Disease. Application to Parkinson’s disease. Row 1 (panels a–c) shows zero-shot, cross-lingual evaluation on NeuroVoz (107 participants, Spanish speech; 5-seed mean ± SD throughout). a, ROC curves for VoiceFM-Whisper (frozen) by task category. The pooled all-tasks ROC is essentially the speech ROC, consistent with a speech-dominated articulatory signal. b, Per-group classification AUROC: 5-fold cross-validated regularized logistic-regression probe trained on each eGeMAPSv02 feature family alone, predicting the PD label; error bars are across-fold standard deviation; the dashed line marks the AUROC of the full-feature LR. Feature counts per group: F0 = 10, Jitter/Shimmer = 4, HNR = 2, Loudness = 12, Formant Amplitudes = 6 (F1–F3 amplitudes relative to F0), Formants = 12 (F1–F3 frequency + bandwidth), Spectral = 21, MFCC = 16, Voicing = 5. c, Cohen’s d effect sizes (PD vs HC) for top 12 eGeMAPSv02 features. Bars are color-coded by feature class — articulatory (red), spectral (orange), voicing (purple), loudness (blue), phonatory (gray) — with the legend at the lower right of the panel. Row 2 (panels d–f) shows fine-tuned evaluation on the mPower sustained-vowel cohort (VoiceFM-Whisper fine-tune; 585 test participants, 101 PD / 484 control). d, Test-set ROC (sustained vs countdown vs combined). e, P(PD) trajectories over the first months of participant enrollment (50 PD vs 50 control participants; thin lines: individual trajectories, thick lines: monthly group means). PD participants retain ≥ 20 contributors per monthly bin through month 5, whereas controls fall below this threshold after month 3; group-mean curves are plotted only up to the point where each group remains adequately sampled. f, Per-group classification AUROC for mPower sustained vowels, same axes and protocol as panel b for visual comparison; feature counts per group are identical to those listed for panel b. g, Cohen’s d for top 12 mPower eGeMAPSv02 features; formant frequencies and bandwidths dominate. Features marked † are sex-confounded and not significant within sex. The sign of Cohen’s d (PD − HC) indicates direction: positive = higher in PD, negative = higher in HC.

B2AI-Voice is predominantly English-language; fewer than 1% of training participants recorded in Spanish, providing minimal Spanish exposure during pre-training. To test whether the learned representations capture language-invariant features of Parkinsonian voice, we evaluated on NeuroVoz [23], a fully Spanish-language PD dataset with 107 participants (52 PD, 55 controls). NeuroVoz includes 2,168 recordings with tasks ranging from sentence reading to diadochokinesis. We extracted VoiceFM-Whisper embeddings with no fine-tuning or language adaptation. We then trained simple logistic regression probes via 5-fold participant-level cross-validation. The model achieved a participant-level AUROC of **0.927 ± 0.015** (**Figure 6a**). This represents strong cross-lingual PD discrimination in a language and acoustic environment far outside the pre-training distribution. These results support the broader hypothesis behind VoiceFM that data-driven contrastive alignment across disease categories produces representations useful across multiple pathologies. They also demonstrate, in this case, that a multi-condition voice foundation model trained on a primarily English clinical cohort generalized to non-English PD detection without retraining.

The transfer was task-dependent in a way that mirrored our recording attribution analysis (**Figure 5a**). For example, we observed highest AUROC (0.933 ± 0.017) on connected sentence reading, followed by diadochokinesis (0.862 ± 0.060), with sustained vowels weakest (0.646 ± 0.033). Tasks that engage articulatory motor control transferred best. Isolated vocal fold tasks transferred worst. This is consistent with the clinical picture of hypokinetic dysarthria in PD as a deficit of speech movement rather than phonation. Notably, the pooled all-tasks AUROC (0.93) was no higher than speech alone. Thus, adding sustained-vowel and DDK recordings did not improve participant-level discrimination, consistent with the speech-dominated articulatory signal.

We then sought to assess whether the cross-lingual signal primarily associates with known acoustic biomarkers. We extracted 88 eGeMAPSv02 features [24] using openSMILE [25] from each NeuroVoz recording. A full 88-feature logistic regression classifier achieved AUROC 0.90, only modestly below VoiceFM’s 0.93. The two approaches’ output probabilities were significantly correlated (Pearson r = 0.62, Spearman ρ = 0.71). Traditional acoustic features therefore captured roughly half of the rank-order variance VoiceFM used for PD discrimination. For each eGeMAPSv02 feature family (e.g., formant amplitudes, voicing, spectral), we then trained a separate logistic-regression classifier on PD vs control with 5-fold cross-validation (**Figure 6b**). The strongest single-family signal came from formant amplitudes — the six F1–F3 amplitude features measured relative to F0. On their own, they reached AUROC 0.91 and produced the largest permutation-importance drop when removed from the full classifier (Δ = −0.12). Voicing and spectral families followed at 0.90 and 0.85. Traditional phonatory markers (jitter, shimmer) were essentially at chance alone (AUROC 0.53). Thus, on clean laboratory recordings, traditional acoustic analysis recovered most — but not all — of the information VoiceFM extracted. The model captured comparable structure from raw audio with no feature engineering. It also generalized across languages without retraining.

By Cohen’s d (**Figure 6c**), the largest PD-vs-control separations were in articulatory features (formant amplitudes, MFCC variability). Traditional phonatory markers (jitter, shimmer, HNR) showed small effects. These results contradict the conventional clinical voice biomarker framing that emphasizes phonatory measures (jitter and shimmer). They instead support an articulatory-deficit account of Parkinsonian dysarthria, one the model discovered without explicit acoustic-feature targets.

Having established cross-lingual transfer on clean laboratory audio, we next asked whether VoiceFM also generalized to large-scale, real-world smartphone recordings. We fine-tuned VoiceFM-Whisper on the mPower Parkinson’s study [26], a longitudinal smartphone-based cohort with 5,044 participants (970 PD, 4,074 controls) and 126,510 sustained phonation and countdown recordings. The fine-tuned model achieved participant-level AUROC of 0.870 on 585 held-out test participants (101 PD, 484 controls), with sustained vowels reaching 0.854 and the countdown task 0.802 (**Figure 6d**). Each participant’s probability of PD was computed as the mean of all their recording-level predictions. Recent literature reports a wide range of participant-level mPower AUROCs depending on cohort curation, from approximately 0.7 in early work [27] to 0.94–0.96 in recent deep-learning studies that apply stringent age and recording-quality filtering [28,29]. Our 0.87 sat within this range. It was achieved on the full 585-participant test cohort without age curation, using a model pre-trained on a multi-disease cohort rather than a model purpose-built for PD detection.

Since mPower has recordings obtained at multiple time points for a subset of participants, we asked whether VoiceFM-Whisper detected a stable trait or a fluctuating state. We found that predictions remained relatively stable over the participant-enrollment window where coverage permitted monthly estimates (**Figure 6e**). Individual-level P(PD) trajectories for 50 PD and 50 control participants separated cleanly, with monthly group means near 0.7 for PD and 0.2 for controls. Paired within-participant analysis of recordings made before versus after levodopa medication showed no significant difference in P(PD) (mean delta = +0.001, p = 0.79). Our interpretation is that the model reliably detects the underlying condition, and not just acute motor symptoms.

Here as well, we sought to assess whether VoiceFM captures traditional acoustic signal from the cell phone recordings (**Figure 6f**). We found that a full 88-eGeMAPSv02 feature logistic regression reached AUROC 0.71, which is below VoiceFM-Whisper’s fine-tuned 0.87. Per-participant agreement was correspondingly lower (Pearson r = 0.32). No single family dominated on mPower. Loudness, spectral, F0, and formant frequency/bandwidth all contributed roughly equally (alone AUROC ≈ 0.6–0.7). Formant amplitudes, the dominant family on clean NeuroVoz audio (AUROC 0.91), collapsed to chance level (AUROC 0.54), consistent with amplitude-based measures being sensitive to recording conditions. By Cohen’s d (**Figure 6g**), the largest PD-vs-control differences on mPower were again articulatory and prosodic (F2 bandwidth variability, F2/F3 frequency, F0 mean). Phonatory markers were consistently uninformative across both cohorts.

## Discussion

We have shown here that contrastive alignment with clinical metadata produces voice representations that transfer across diseases, languages, and recording conditions. We tested three audio backbones spanning an order of magnitude in scale and three very different pre-training objectives: Whisper, trained for multilingual speech recognition; HuBERT, trained with masked-unit prediction on English speech; and HeAR, trained with masked autoencoding on health-related audio. The contrastive strategy improved all three. The gains therefore reflect the alignment objective itself, not the size or architecture of any one encoder. We use the term “foundation model” in a directional sense: VoiceFM was trained on far fewer participants than the million-sample datasets that define vision or language foundation models, and whether this strategy scales with more participants remains an open empirical question.

Our results suggest that contrastive learning produces strong clinical voice representations. Our best model, VoiceFM-Whisper, achieved mean AUROC 0.952 across the five evaluation tasks (four disease categories plus control vs disease screening). This was done using only task-agnostic contrastive training with linear probes. VoiceFM-Whisper significantly outperformed a contrastively-trained HuBERT counterpart as well as the two models’ frozen baselines (Whisper-v2 and HuBERT). We hypothesize that Whisper’s larger representation (1280 dimensions, 32 layers trained on 680,000 hours of multilingual audio) provides a substantially stronger foundation than HuBERT (768 dimensions, 12 layers, LibriSpeech only), and contrastive fine-tuning amplified this advantage. The contrastive approach yields a general-purpose embedding that can be probed for any downstream task, including tasks not seen during training, without retraining the model. Importantly, the temporally held-out evaluation (**Figure 3**) showed that this performance generalized to participants enrolled after model freeze at the same sites. The distinct test of distribution shift across sites and recording conditions was provided by external datasets including NeuroVoz and mPower.

The B2AI-Voice protocol comprises 22 standardized speech and voice task families per participant; because each family includes several prompt variants, the dataset contains ∼65 unique recording types after participant-coverage filtering. We asked whether a smaller subset of recording types would suffice for disease classification. Our task attribution analysis suggests it would: a curated subset of about ten recording types on average matched or slightly exceeded the full set’s performance. A targeted protocol could therefore in theory replace a lengthy comprehensive session while improving both diagnostic performance and patient compliance.

Our cross-lingual transfer results on NeuroVoz suggest that VoiceFM-Whisper embeddings generalize beyond English without any fine-tuning or language-specific adaptation. The acoustic explainability contrast between NeuroVoz and mPower suggests that recording quality alters what the model relies on: on clean lab recordings, standard eGeMAPSv02 features approached VoiceFM’s discrimination. On noisy smartphone data the gap widened substantially and a sizable share of the model’s signal lay beyond what traditional hand-crafted acoustic features capture.

In both NeuroVoz and mPower PD cohorts, our acoustic decomposition analyses consistently implicate articulatory rather than phonatory features. Formant amplitudes relative to F0, formant bandwidth variability, and MFCC dynamics dominated the PD detection signal. At the same time, traditional phonatory markers (jitter, shimmer, HNR) contributed minimally. This pattern is consistent with the clinical picture of hypokinetic dysarthria. Accordingly, the recording task attribution results showed that speech tasks outperformed sustained phonation tasks. These results also align with the segment-dependent articulatory framing reported by Williamson et al. on the Interspeech 2015 ComParE Parkinson’s challenge [30], where speech-segment dynamics (not phonation-derived stability measures) carried most of the UPDRS-prediction signal.

Our study has several limitations. First, the B2AI-Voice dataset, while multi-site, contains 984 participants meeting our labeling criteria (846 training plus 138 temporally held out). While large by the standards of the audiomics field, its limited size may not enable enough statistical power for rare conditions. Limited size may also make results sensitive to train/test splits, as evidenced by the inter-seed variance in mood disorder and COPD/asthma classification.

Finally, we also noted that the dominant Cohen’s d features on both NeuroVoz (formant amplitudes) and mPower (formant frequencies and bandwidth variability) are articulatory. As a further check, we computed Cohen’s d effect sizes separately within male and within female mPower participants (data not shown). The strongest articulatory features (formant frequencies and bandwidth variability) still showed large PD-vs-control separations within each sex, indicating that these features reflect disease rather than sex differences between the PD and control groups.A third limitation concerns output calibration. We did not explicitly assess the calibration of our classifiers’ probability outputs, and they should not be interpreted as true posterior probabilities without a post-hoc calibration step (e.g., Platt scaling) on a representative validation set. The longitudinal trajectory analysis on mPower interprets relative trends in P(PD) over time rather than absolute probability values, which limits the impact of miscalibration on those conclusions. Clinical deployment of any of these models would require explicit calibration.

A fourth limitation is generalizability: while the NeuroVoz results suggest cross-lingual transfer to Spanish, generalization to more distant languages and noisier recording conditions remains untested. In general, scaling VoiceFM to larger and more diverse datasets will be needed to improve performance, especially for some conditions such as respiratory diseases where B2AI-Voice data is limited. We envision that combining B2AI-Voice with public voice datasets might produce models with broader clinical coverage and predictive capabilities. The B2AI-Voice dataset does not have any longitudinal voice collection. Longitudinal voice assessment could test whether embeddings track disease progression or treatment response across conditions. Perceptual speech outcomes such as speech intelligibility (also not currently captured in B2AI-Voice) represent another area for further work, and would require structured elicitation (e.g., a standardized reading passage scored by listeners or automated intelligibility metrics). Another direction for further research involves federated or privacy-preserving training approaches. These would address the legitimate concerns about voice biometric data while further enabling multi-institutional collaboration. On the clinical-encoder side, replacing the TabTransformer with newer tabular foundation models such as TabICL or TabDPT may offer further gains as the clinical phenotype space grows. Despite all these limitations and opportunities for further research, taken together, the results presented in this manuscript provide evidence that foundation model approaches can advance voice biomarker research beyond single-condition, single-dataset paradigms toward general-purpose, multi-condition voice-based screening.

## Methods

### Dataset: Bridge2AI-Voice as a Biomarker of Health

The Bridge2AI-Voice as a Biomarker of Health (B2AI-Voice) dataset was collected across five academic medical centers: University of South Florida (USF, n = 426 in training cohort), Sinai Health Systems (n = 160), Vanderbilt University Medical Center (VUMC, n = 152), Weill Cornell Medicine (WCM, n = 64), and Massachusetts Institute of Technology (MIT, n = 44) [18]. Participants were recruited from disease-specific clinical populations (voice disorders, neurodegenerative disease, mood and respiratory disorders) and controls. For the adult cohort, inclusion required age ≥ 18, ability to consent, and English or Spanish fluency. Audio was recorded in clinical or quiet environments using standardized equipment at each site. The equipment at each site included an iPad for the recording interface and an AVID-Headset, which participants wore to stabilize microphone-to-mouth distance, following the Bridge2AI-Voice recording protocol [31].

Clinical metadata was collected via REDCap and included demographics (age, sex, ethnicity, race), validated clinical instruments (PHQ-9 for depression, GAD-7 for anxiety, VHI-10 for voice handicap), disease category flags (self-reported and clinician-validated GSD), and clinician-rated perceptual voice assessments (Consensus Auditory-Perceptual Evaluation of Voice [CAPE-V]). All analyses in this paper use the most recent clinically curated B2AI-Voice data release and gold-standard diagnosis (GSD) labels unless otherwise specified. The GSD labeling protocol defines a control as a participant without any of the clinical diagnoses specifically studied in this cohort and without an abnormal-sounding voice on review by the research assistant or clinician (**Table S2** lists per-diagnosis cohort sizes). GSD flags can co-occur at both the diagnosis and category level: 85 participants carry two or more GSD diagnosis flags, and 60 participants meet criteria for two or more broader disease categories (voice, neurological, mood, respiratory). Participants who could not be assigned to either control or any disease category under this protocol (n = 25) were excluded from all analyses.

VoiceFM was trained on the 846 participants available at the time of model development; an additional 138 participants were enrolled in B2AI-Voice after training was finalized, at the same five sites and under the same protocol, and used only for temporally held-out inference (**Figure 3**). Within the training cohort, splits were 70/15/15 (train/validation/test) by participant, stratified by disease category, using seed 42; multi-seed evaluation used seeds 42–46.

### Audio preprocessing

We resampled all audio files to 16 kHz mono. Recordings were trimmed to a maximum of 30 seconds, with leading and trailing silence removed using a −40 dB threshold. Amplitude was normalized to unit peak. Recording durations varied substantially across task types (mean 15.8 s, median 8.4 s, IQR 4.6–20.7 s, range 0.1–332.5 s; 176 total hours of raw audio across the 984-participant cohort). For the mPower dataset, m4a files (Advanced Audio Coding [AAC] codec) were batch-converted to wav using ffmpeg prior to training.

### VoiceFM model architecture

VoiceFM uses a CLIP-style [19] dual-encoder architecture. We evaluated two audio backbones. The primary model uses the Whisper large-v2 encoder [22] (32 transformer layers, 1280 hidden dimensions, ∼637M parameters; the full Whisper large-v2 model has ∼1.55B parameters but only the encoder is used here). The encoder weights were initialized from the public Whisper large-v2 checkpoint pretrained on 680,000 hours of multilingual audio. Layers 0–27 are frozen; layers 28–31 are fine-tuned. Frame-level encoder outputs are aggregated via mean pooling and projected to 256 dimensions via a linear layer with LayerNorm and Gaussian Error Linear Unit (GELU) activation. Whisper produces a fixed 1500-frame temporal grid for any input, which removed the variable-length motivation for the attentive pool we used on HuBERT. The secondary model uses HuBERT-base [15] (12 transformer layers, 768 hidden dimensions, 95M parameters pretrained on LibriSpeech). Layers 0–8 are frozen; layers 9–11 are fine-tuned (this depth was selected by validation retrieval R@5 among configurations unfreezing 3, 4, or 6 layers; the Whisper freeze depth was set proportionally and not separately ablated). Frame-level HuBERT outputs are aggregated via attentive pooling (learned query vector attending over frames) and projected to 256 dimensions. As an additional baseline, we evaluated Google’s Health Acoustic Representations (HeAR) model [17], a ViT-L encoder (303M parameters) pretrained on health-related audio via masked autoencoding. HeAR accepts fixed 2-second audio inputs; we split each recording into non-overlapping 2-second chunks, extracted 512-dimensional embeddings from each chunk using the frozen encoder, mean-pooled across chunks, and projected to 256 dimensions. Only the projection layer was trainable (4.1M parameters).

The clinical encoder is a tabular transformer [20] with 4 layers, 4 attention heads, and 256 hidden dimensions. Input features (44 total: 36 binary, 4 continuous, 4 categorical; **Table S1**) are tokenized into learned embeddings, with continuous features discretized into bins. The transformer output is mean-pooled and projected to the shared 256-dimensional embedding space.

Training minimizes symmetric InfoNCE loss between audio and clinical embeddings:

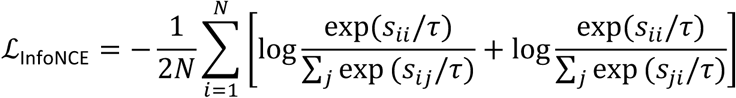

where *s_ij_* is the cosine similarity between audio embedding *i* and clinical embedding *j*, and *τ* is a learned temperature parameter. Auxiliary losses include disease category prediction (binary cross-entropy, weight 0.1) and age regression (MSE, weight 0.05).

Training used AdamW optimizer with learning rates of 1×10⁻⁵ (backbone) and 1×10⁻⁴ (projection heads), cosine annealing with 10% warmup, effective batch size 64 (batch 32 × gradient accumulation 2), and hard negative mining (beta = 1.0). Models were trained for up to 200 epochs with early stopping (patience 25) on validation loss. During training, one random recording per participant is sampled each epoch, ensuring each participant contributes equally regardless of recording count.

### Gold-standard diagnosis classification

We trained and evaluated four model configurations across 5 random seeds (42–46), each with different train/val/test splits stratified by disease category. All models were evaluated using identical methodology (same evaluation script, same splits, same logistic regression probe hyperparameters):

1. **VoiceFM-Whisper (contrastive):** Frozen 256-dimensional audio embeddings from VoiceFM with the Whisper large-v2 backbone, evaluated via logistic regression probes.
2. **VoiceFM-HuBERT (contrastive):** Frozen 256-dimensional audio embeddings from VoiceFM with the HuBERT-base backbone, evaluated via logistic regression probes.
3. **Frozen Whisper baseline:** Mean-pooled 1280-dimensional embeddings from unmodified Whisper large-v2 encoder, evaluated via logistic regression probes.
4. **Frozen HuBERT baseline:** Mean-pooled 768-dimensional embeddings from unmodified HuBERT-base, evaluated via logistic regression probes.

Evaluation metrics: AUROC per disease category (control vs disease, voice, neurological, mood, respiratory), computed at participant level (one embedding per participant, mean-pooled across recordings). Per-diagnosis AUROC was computed for individual GSD conditions with N ≥ 10 participants.

### Evaluation on the temporally held-out cohort

We assembled the 138 temporally held-out participants (38 controls, 46 voice, 23 neurological, 12 mood, 31 respiratory) by retaining all participants in the most recent B2AI-Voice release that were not part of the original training cohort and that satisfied the same GSD labeling protocol. They were enrolled at the same five academic medical centers and under the same recording protocol as the training cohort, so the cohort tests temporal generalization to new patients within the original collection environment rather than distribution shift across sites or instruments. For each disease category and individual diagnosis, we trained logistic regression probes on participant-mean VoiceFM-Whisper embeddings restricted to the training cohort, and applied the resulting probe to the held-out 138 participants without any retraining or fine-tuning. Five random seeds (42–46) were averaged for the held-out AUROCs reported in **Figure 3** (5-seed mean ± SD); the held-out cohort is fixed across seeds, so the reported variance reflects probe sensitivity to the training-cohort split rather than additional resampling of the validation cohort. The held-out evaluation uses the same probe pipeline and the same StandardScaler-fit-on-training, evaluate-on-held-out convention as the standard cross-validation experiments.

### External dataset transfer

We evaluated frozen VoiceFM-Whisper and Frozen Whisper embeddings on three external voice datasets via 5-fold stratified cross-validation with logistic regression:

- **Coswara** [32]: COVID-19 detection from cough/breathing/speech (n = 2,098)
- **Saarbrücken Voice Database (SVD)** [33]: Voice pathology detection from sustained vowels (n = 2,041)
- **MDVR-KCL** [34]: Parkinson’s detection from reading passages (n = 73)

Few-shot evaluation: for each external dataset and each k ∈ {1, 2, 5, 10, 20}, we randomly sampled k participants per class to serve as the training set, fit a logistic-regression probe on their frozen 256-d VoiceFM-Whisper (or 1280-d Frozen Whisper) embeddings standardized with a per-fit StandardScaler, and evaluated participant-level AUROC on the remaining participants. We repeated this 100 times per k and report the mean ± standard deviation across trials. The audio encoder is never updated; only the linear probe is fit on each draw.

### Recording type attribution

We performed greedy forward selection of recording types. Starting from no recordings, we iteratively added the recording type that most increased mean AUROC across all evaluable disease categories, with embeddings mean-pooled per participant and evaluated via logistic regression. We repeated the procedure for five seeds (42–46) to assess stability of the selected types.

For the per-recording-type evaluation, we trained and evaluated an independent logistic-regression probe for each recording type that had at least 20 test participants per disease class, reporting AUROC per disease category. Between 65 and 67 recording types met this threshold per seed, depending on the train/test split.

As a selection-bias sensitivity check, we ran a nested-cross-validation variant of the greedy procedure. Because the primary analysis selects subsets directly on the test split, absolute peak AUROC values can be optimistically biased. In the nested variant we used the 15% held-out validation fold as the selection signal: at each greedy step we added the candidate recording type that most increased mean validation AUROC, while test AUROC was computed only for record-keeping. We took the cumulative selection at the validation-peak step as the final reported subset. Across five seeds (42–46), the validation-selected subset’s mean test AUROC matched the all-types baseline within seed variability (data not shown), supporting the interpretation that targeted protocols may match — but do not robustly exceed — the full battery.

### NeuroVoz cross-lingual PD detection

#### Dataset

The NeuroVoz dataset [23] contains 2,168 Spanish-language recordings from 107 participants (52 PD, 55 controls) collected under controlled laboratory conditions. Recording types include sentence reading, sustained vowels, and diadochokinesis (pa-ta-ka). PD diagnosis labels were used as ground truth; severity metadata available in the dataset (UPDRS, Hoehn-Yahr) was not used in the analyses reported here.

#### Evaluation

Frozen VoiceFM-Whisper and Frozen Whisper embeddings were extracted from all recordings and mean-pooled per participant. Participant-level PD classification was evaluated via 5-fold stratified cross-validation with logistic regression, repeated across 5 random seeds (42–46). Per-task-category evaluation was performed separately for speech, vowel, and DDK recordings.

#### Acoustic feature analysis

We extracted 88 eGeMAPSv02 features (openSMILE) from all recordings and aggregated per participant. Per-group classification AUROCs (5-fold stratified cross-validated regularized logistic regression per eGeMAPSv02 feature family alone, plus full-feature baseline), joint group permutation importance (30 permutations × 5 folds, shuffling each group’s columns jointly to preserve within-group correlation), Pearson agreement between the eGeMAPSv02-LR out-of-fold probabilities and VoiceFM P(PD), and Cohen’s d effect sizes were computed as described for mPower.

### mPower Parkinson’s disease detection

#### Dataset

The mPower Parkinson’s Disease study [26] was accessed through Sage Bionetworks (Synapse syn4993293). The dataset contains 63,255 sustained phonation recordings and 63,255 countdown recordings (126,510 total) from 5,044 participants (970 self-reported as having a clinician diagnosis of PD, 4,074 controls), collected via smartphone app. Participant-level splits were 70/15/15 (train/val/test), stratified by PD status.

#### Fine-tuning architecture

Whisper large-v2 audio encoder (freeze layers 0–27, fine-tune 28–31, matching VoiceFM-Whisper pre-training), followed by mean pooling, projection to 256 dimensions, and a binary classification head (sigmoid), initialized from VoiceFM-Whisper pre-trained weights. Training used binary cross-entropy with inverse-frequency class weights, AdamW (lr = 1×10⁻⁵ backbone, 1×10⁻⁴ head), cosine annealing, and effective batch size 64. The fine-tune reported in this paper was run for one seed (seed 43) and 5 training epochs, with the best validation epoch (epoch 4, validation participant-level AUROC 0.94) used for test-set evaluation; a full multi-seed × 30-epoch protocol is computationally constrained and reserved for future work.

#### Evaluation

We report participant-level AUROC on the 585-participant held-out test set. Each participant’s P(PD) is the mean of all their recording-level sigmoid outputs. Per-task type AUROCs (sustained vs countdown) were recovered from the dataset-filtered metadata order using positional index. Threshold optimization used Youden’s J statistic on the validation set.

#### Longitudinal trajectory analysis

For participants with ≥ 2 sustained vowel recordings, P(PD) was computed per recording and aggregated into monthly bins. Trajectories were truncated at 5 months and required ≥ 20 participants per bin for stable estimates.

#### Acoustic feature analysis

For both NeuroVoz and mPower, we extracted 88 eGeMAPSv02 features [24] using openSMILE on every test-set sustained vowel recording, aggregated to participant level by mean-pooling, and partitioned them into nine non-overlapping families assigned by strict priority (F0, jitter/shimmer, HNR, loudness, formant amplitudes, formant frequency + bandwidth, spectral, MFCC, voicing). For each family we report two quantities. First, the 5-fold stratified cross-validated AUROC of a regularized logistic-regression probe trained on that family alone, with StandardScaler fit inside each training fold to avoid leakage. Second, the AUROC drop from joint-permutation importance: at each fold we fit the full 88-feature LR on the training split, then for each family we applied a single random permutation index to all of that family’s columns of the held-out fold (30 permutations averaged), preserving within-family correlation. To quantify cross-system agreement we additionally compute Pearson r between the eGeMAPSv02-LR out-of-fold probabilities and VoiceFM P(PD) on the same participants. Cohen’s d effect sizes between PD and control participants were computed per feature using a pooled standard deviation; features were ranked by |d|. The mPower analysis used all 15,166 sustained recordings from the 585 test participants (101 PD, 484 controls); the NeuroVoz analysis used the full 107-participant cohort.

### Embedding interpretability

#### Acoustic grounding

We extracted 14 Praat/parselmouth features (F0 mean/SD, jitter, shimmer, HNR, CPPS, formants F1–F3) from prolonged vowel recordings (878 participants) and trained ridge regression probes (alpha = 1.0, StandardScaler) with 5-fold cross-validation across all participants; the acoustic features were not used during model training. We compared two embedding types: VoiceFM-Whisper (256d) and Frozen Whisper (1280d). We then ran PCA followed by Spearman rank correlation between the acoustic features and the top principal components.

#### Nearest-neighbor retrieval

For each participant, we retrieved the k = 5 nearest neighbors in VoiceFM-Whisper embedding space (cosine distance) and measured clinical similarity: diagnosis category match rate, severity score difference (CAPE-V), and voice quality difference (self-rated 1–10).

#### Within-participant consistency

We computed the cosine similarity between a participant’s VoiceFM-Whisper embeddings from different recording types (intra-person similarity) and between different participants’ embeddings from the same recording type (inter-person similarity). The separation between these two distributions indicates the strength of participant-level identity encoding.

#### CKA layer analysis

We computed linear centered kernel alignment (CKA) [35] between corresponding layers of Frozen Whisper large-v2 and VoiceFM-Whisper’s fine-tuned Whisper encoder on 500 prolonged vowel recordings, across all 32 transformer layers. CKA ranges from 0 (completely different representations) to 1 (identical).

### Demographic-shortcut audit

To assess whether VoiceFM’s classification performance reduces to a demographic shortcut, we ran two complementary analyses on the training cohort. (i) Demographic-only baseline: a logistic regression classifier on age and binary sex (is_male) was trained for each disease category using 5 random seeds × 5-fold StratifiedKFold cross-validation, with StandardScaler applied within fold. (ii) Residualization: we regressed each VoiceFM-Whisper embedding dimension on age and sex (linear regression on the full cohort), took residuals, and re-ran the same logistic-regression probe. The change in AUROC between the original embeddings and the residualized embeddings (Δ AUROC) quantifies the demographic dependence of each category.

### Statistical analysis

We report all classification performance as AUROC with participant-level aggregation (mean probability across recordings per participant). Five-seed experiments report mean ± standard deviation. Few-shot experiments report mean ± standard deviation across 100 random trials. Significance testing uses two-sided Welch’s t-tests (unpaired, unequal variance) for model comparisons across seeds. Mann-Whitney U tests are used for unpaired group comparisons.

### Reporting standards

We report results in alignment with relevant items of the TRIPOD-AI (Transparent Reporting of a multivariable prediction model for Individual Prognosis Or Diagnosis — Artificial Intelligence) 2024 reporting guidance for prediction model research [36]. The TRIPOD-AI items are addressed across the manuscript as follows:

- Participant characteristics (training and temporally held-out cohorts): **Tables 1 and 2**
- GSD labeling exclusion criteria: **Methods**
- Model development and validation: subsections “Gold-standard diagnosis classification”, “Evaluation on the temporally held-out cohort”, “External dataset transfer”, “NeuroVoz cross-lingual PD detection”, and “mPower Parkinson’s disease detection”
- Performance metrics (AUROC with uncertainty estimates, Cohen’s d): **Results** and **Figures 2–6**
- Model interpretability and limitations: **Discussion** Participant-level cross-validation, temporally held-out evaluation, and external-dataset evaluation prevent information leakage across training and evaluation. The full source code, configurations, and figure-generation scripts are available at https://github.com/oelemento/VoiceFM-public.

## Author contributions

O.E. and A.R. conceived the project and designed the study. O.E. designed and implemented the VoiceFM model, training pipeline, and evaluation framework, and led the experiments and statistical analyses. A.S. contributed to data preparation and evaluation-framework implementation. J.T.C. and S.S.G. contributed to acoustic-feature analyses and external-dataset evaluation. I.H. contributed to data analysis and manuscript preparation. Y.B. and the Bridge2AI-Voice Consortium designed and executed the Bridge2AI-Voice data collection protocol and provided clinical phenotyping. O.E. drafted the manuscript with input from all authors. All authors reviewed and approved the final manuscript.

## AI use disclosure

The authors used Claude (Anthropic) for manuscript drafting and copy-editing assistance during the preparation of this manuscript. All scientific claims, analyses, interpretations, and final wording are the authors’ own, and the authors take full responsibility for the content of the manuscript. No AI tool was used to generate analysis code that produced reportable results without author review, or to create or edit figures containing research results.

## Data availability

The Bridge2AI-Voice dataset is distributed via PhysioNet and Sage Bionetworks (https://doi.org/10.13026/k81f-qr68) [18] subject to a data-use agreement. The mPower Parkinson’s Disease study data are available via Sage Bionetworks (Synapse syn4993293) under the mPower data access terms. NeuroVoz [23], MDVR-KCL [34], Saarbrücken Voice Database [33], and Coswara [32] are publicly available from their respective sources cited in the References.

## Code availability

The full source code, configurations, and figure-generation scripts used in this study are available at https://github.com/oelemento/VoiceFM-public under the MIT License. Reproducing the primary figures requires only the result JSONs distributed with the repository; reproducing model training requires access to the Bridge2AI-Voice dataset under the data-use agreement above.

## Bridge2AI-Voice Consortium members

The following individuals are members of the Bridge2AI-Voice Consortium and contributed to the design, recruitment, data collection, ethical oversight, and infrastructure of the Bridge2AI-Voice study (named co-authors above are not re-listed here):

Maria Powell (Vanderbilt University Medical Center, Nashville, TN, USA); David Dorr (Oregon Health & Science University, Portland, OR, USA); Phillip Payne (Washington University in St. Louis School of Medicine, St. Louis, MO, USA); Vardit Ravitsky (The Hastings Center, Garrison, NY, USA); Jean-Christophe Bélisle-Pipon (Simon Fraser University, Burnaby, BC, Canada); Ruth Bahr (University of South Florida, Tampa, FL, USA); Stephanie Watts (University of South Florida, Tampa, FL, USA); Donald Bolser (University of Florida, Gainesville, FL); Jennifer Siu (Hospital for Sick Children, Toronto, ON, Canada); Jordan Lerner-Ellis (University of Toronto, Toronto, ON, Canada); Frank Rudzicz (Dalhousie University; Vector Institute, Halifax, NS, Canada); Micah Boyer (University of South Florida, Tampa, FL, USA); Yassmeen Abdel-Aty (University of South Florida, Tampa, FL, USA); Toufeeq Ahmed Syed (UTHealth Houston, Houston, TX, USA); James Anibal (NIH Clinical Center, U.S. National Institutes of Health; Institute of Biomedical Engineering, University of Oxford, Bethesda, Maryland, USA); Dona Amraei (Simon Fraser University, Burnaby, BC, Canada); Stephen Aradi (University of South Florida, Tampa, FL, USA); Kirollos Armosh (University of South Florida, Tampa, FL, USA); Ana Sophia Avila Martinez (University of South Florida, Tampa, FL, USA); Shaheen Awan (University of Central Florida, Orlando, FL, USA); Steven Bedrick (Oregon Health & Science University, Portland, OR, USA); Helena Beltran (University of South Florida, Tampa, FL, USA); Alexander Bernier (Simon Fraser University, Burnaby, BC, Canada); Moroni Berrios (University of South Florida, Tampa, FL, USA); Isaac Bevers (Massachusetts Institute of Technology, Cambridge, MA, USA); Alden Blatter (Simon Fraser University, Burnaby, BC, Canada); Rahul Brito (Harvard University; Massachusetts Institute of Technology, Boston, MA, USA); Amy Brown (Vanderbilt University Medical Center, Nashville, TN, USA); John Brown (University of South Florida, Tampa, FL, USA); Léo Cadillac (Simon Fraser University, Burnaby, BC, Canada); Selina Casalino (Mount Sinai Hospital, Sinai Health, Toronto, ON, Canada); Amanda Chao (Baycrest Centre, North York, ON, Canada); John Costello (Boston Children’s Hospital, Boston, MA, USA); Abhijeet Dalal (Oregon Health & Science University, Portland, OR, USA); Iris De Santiago (University of South Florida, Tampa, FL, USA); Enrique Diaz-Ocampo (CENIDET, Cuernavaca, Mexico); Amanda Doherty-Kirby (Simon Fraser University, Burnaby, BC, Canada); Mohamed Ebraheem (University of South Florida, Tampa, FL, USA); Ellie Eiseman (University of South Florida, Tampa, FL, USA); Mahmoud Elmahdy (University of South Florida, Tampa, FL, USA); Renee English (Simon Fraser University, Burnaby, BC, Canada); Emily Evangelista (University of South Florida, Tampa, FL, USA); Kenneth Fletcher (Vanderbilt University Medical Center, Nashville, TN, USA); Hortense Gallois (Simon Fraser University, Burnaby, BC, Canada); C. Gaelyn Garrett (Vanderbilt University Medical Center, Nashville, TN, USA); Alexander Gelbard (Vanderbilt University, Nashville, TN, USA); Omar Ghaffar (Hennick Bridgepoint Hospital, Toronto, ON, Canada); Amer Ghavanini (Trillium Health Partners and University of Toronto, Toronto, ON, Canada); Anna Goldenberg (The Hospital for Sick Children, Toronto, ON, Canada); Karim Hanna (University of South Florida, Tampa, FL, USA); William Hersh (Oregon Health & Science University, Portland, OR, USA); Jennifer Jain (University of South Florida, Tampa, FL, USA); Lochana Jayachandran (Sinai Health, Toronto, ON, Canada); Kaley Jenney (Massachusetts Institute of Technology, Cambridge, MA, USA); Kathy Jenkins (Boston Children’s Hospital, Boston, MA, USA); Stacy Jo (Boston Children’s Hospital, Boston, MA, USA); Alistair Johnson (University of Toronto, Toronto, ON, Canada); Brenda Juan Guardela (University of South Florida, Tampa, FL, USA); Ayush Kalia (University of South Florida, Tampa, FL, USA); Megha Kalia (University of South Florida, Tampa, FL, USA); Zoha Khawaja (Simon Fraser University, Burnaby, BC, Canada); Kenji Kobayashi (Vanderbilt University Medical Center, Nashville, TN, USA); Cynthia Kostelnik (University of South Florida, Tampa, FL, USA); Alisa Krause (University of South Florida, Tampa, FL, USA); Andrea Krussel (Washington University in St. Louis, St. Louis, MO, USA); Elisa Lapadula (Sinai Health, Toronto, ON, Canada); Genelle Leo (University of South Florida, Tampa, FL, USA); Justin Levinsky (Hospital for Sick Children, Toronto, ON, Canada); Chloe Loewith (Simon Fraser University, Burnaby, BC, Canada); Linda Ma (Baycrest Centre, North York, ON, Canada); Radhika Mahajan (Mount Sinai Hospital, Sinai Health, Toronto; Lunenfeld-Tanenbaum Research Institute, Sinai Health, Toronto, Toronto, ON, Canada); Vrishni Maharaj (University of South Florida, Tampa, FL, USA); Marie-Françoise Malo (Simon Fraser University, Burnaby, BC, Canada); Siyu Miao (The Hospital for Sick Children, Toronto, ON, Canada); LeAnn Michaels (Oregon Health & Science University, Portland, OR, USA); Matthew Mifsud (University of South Florida, Tampa, FL, USA); Marian Mikael (University of South Florida, Tampa, FL, USA); Pablo Montoya Varela (Simon Fraser University, Burnaby, BC, Canada); Elijah Moothedan (Florida Atlantic University, Boca Raton, FL, USA); Yosef Nafii (University of South Florida, Tampa, FL, USA); Tempestt Neal (University of South Florida, Tampa, FL, USA); Karlee Newberry (University of South Florida, Tampa, FL, USA); Evan Ng (The Hospital for Sick Children, Toronto, ON, Canada); Christopher Nickel (University of South Florida, Tampa, FL, USA); Amanda Peltier (Vanderbilt University Medical Center, Nashville, TN, USA); Trevor Pharr (University of South Florida, Tampa, FL, USA); Michaela Pnacekova (Simon Fraser University, Burnaby, BC, Canada); Matthew Pontell (Vanderbilt University Medical Center, Nashville, TN, USA); Jaiden Potter (Simon Fraser University, Burnaby, BC, Canada); Claire Premi-Bortolotto (Simon Fraser University, Burnaby, BC, Canada); Parnaz Rafatjou (University of South Florida, Tampa, FL, USA); JM Rahman (University of South Florida, Tampa, FL, USA); Gayathiri Rajkumar (Baycrest Centre, North York, ON, Canada); John Ramos (Weill Cornell Medicine, New York, NY, USA); Sarah Rohde (Vanderbilt University Medical Center, Nashville, TN, USA); Michael de Riesthal (Vanderbilt University Medical Center, Nashville, TN, USA); Jillian Rossi (University of South Florida, Tampa, FL, USA); Laurie Russell (Hospital for Sick Children, Toronto, ON, Canada); Samantha Salvi Cruz (Vanderbilt University Medical Center, Nashville, TN, USA); Joyce Samuel (Hospital for Sick Children, Toronto, ON, Canada); Suketu Shah (University of South Florida, Tampa, FL, USA); Ahmed Shawkat (University of South Florida, Tampa, FL, USA); Elizabeth Silberholz (Boston Children’s Hospital, Boston, MA, USA); John Stark (University of South Florida, Tampa, FL, USA); Lala Su (Hospital for Sick Children, Toronto, ON, Canada); Shrramana Ganesh Sudhakar (University of South Florida, Tampa, FL, USA); Duncan Sutherland (New York, NY, USA); Venkata Swarna Mukhi Talluri (Oregon Health & Science University, Tempe, AZ, USA); Jeffrey Tang (Weill Cornell Medicine, New York, NY, USA); Luka Taylor (Simon Fraser University, Burnaby, BC, Canada); Jamie Toghranegar (University of South Florida, Tampa, FL, USA); Julie Tu (Mount Sinai Hospital, Toronto, ON, Canada); Megan Urbano (University of South Florida, Tampa, FL, USA); Gavin Victor (Simon Fraser University, Burnaby, BC, Canada); Kimberly Vinson (Vanderbilt University Medical Center, Nashville, TN, USA); Jordan Wilke (Massachusetts Institute of Technology, Cambridge, MA, USA); Claire Wilson (Simon Fraser University, Burnaby, BC, Canada); Madeleine Zanin (Mount Sinai Hospital, Toronto, ON, Canada); Xijie Zeng (Dalhousie University & Vector Institute, Halifax, NS, Canada); Theresa Zesiewicz (University of South Florida, Tampa, FL, USA); Robin Zhao (Weill Cornell Medicine, New York, NY, USA); Pantelis Zisimopoulos (Weill Cornell Medicine, New York, NY, USA).

## Supplementary Materials

**Table S1:**
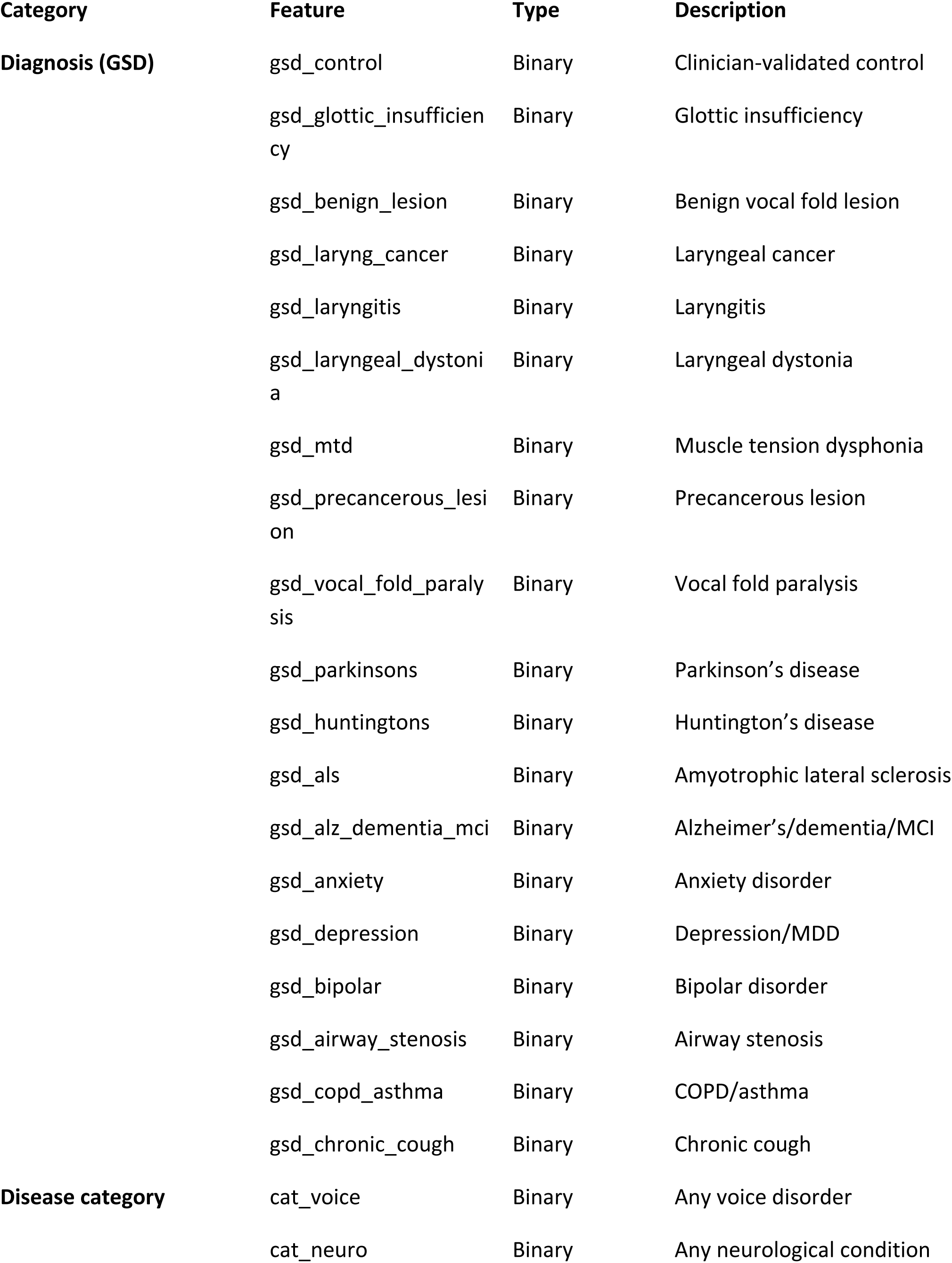
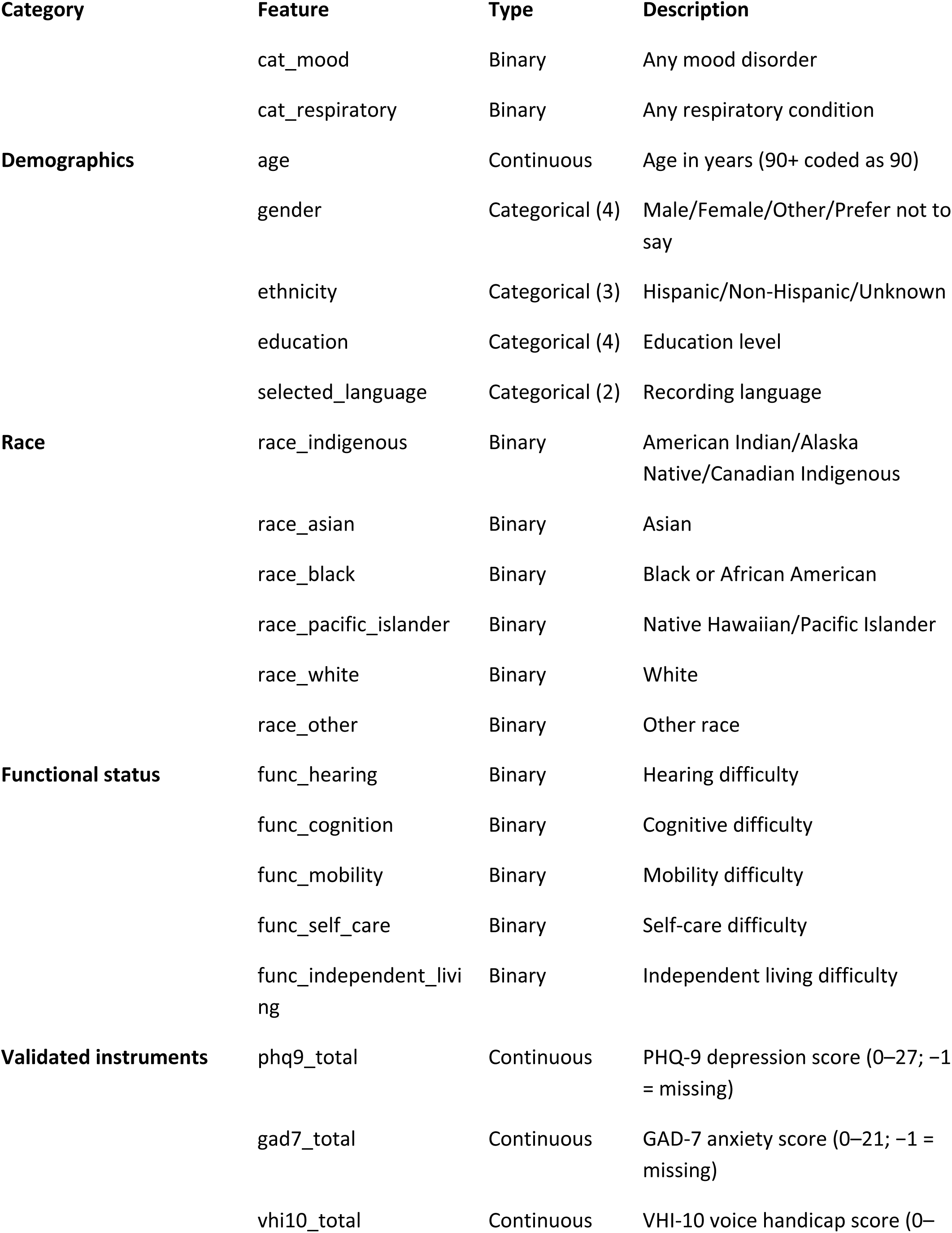
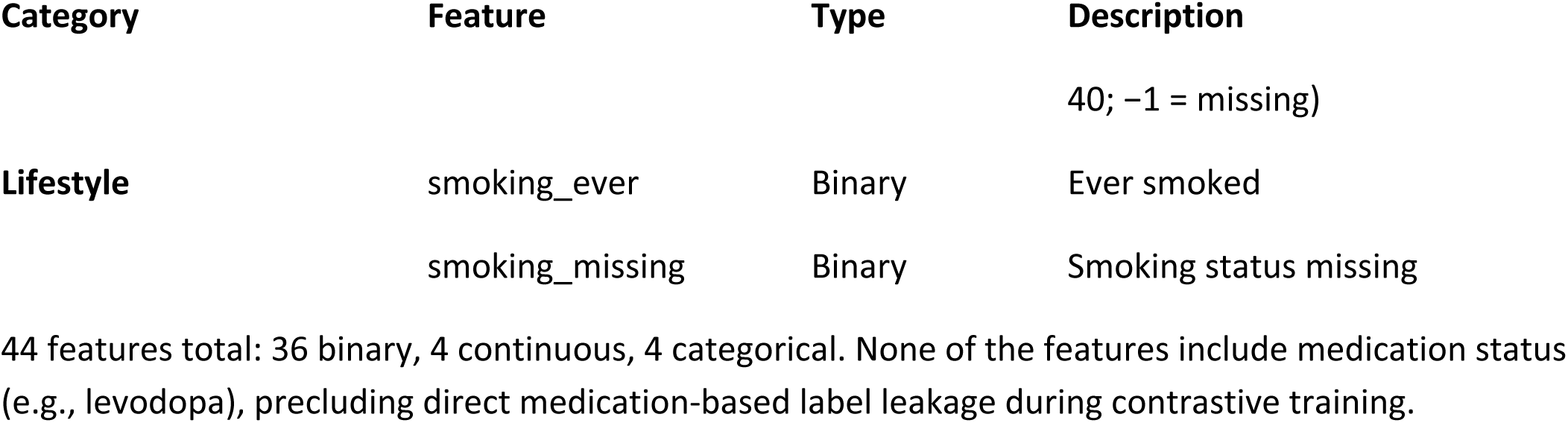
Clinical features used in VoiceFM contrastive training (GSD mode)

**Table S2:**
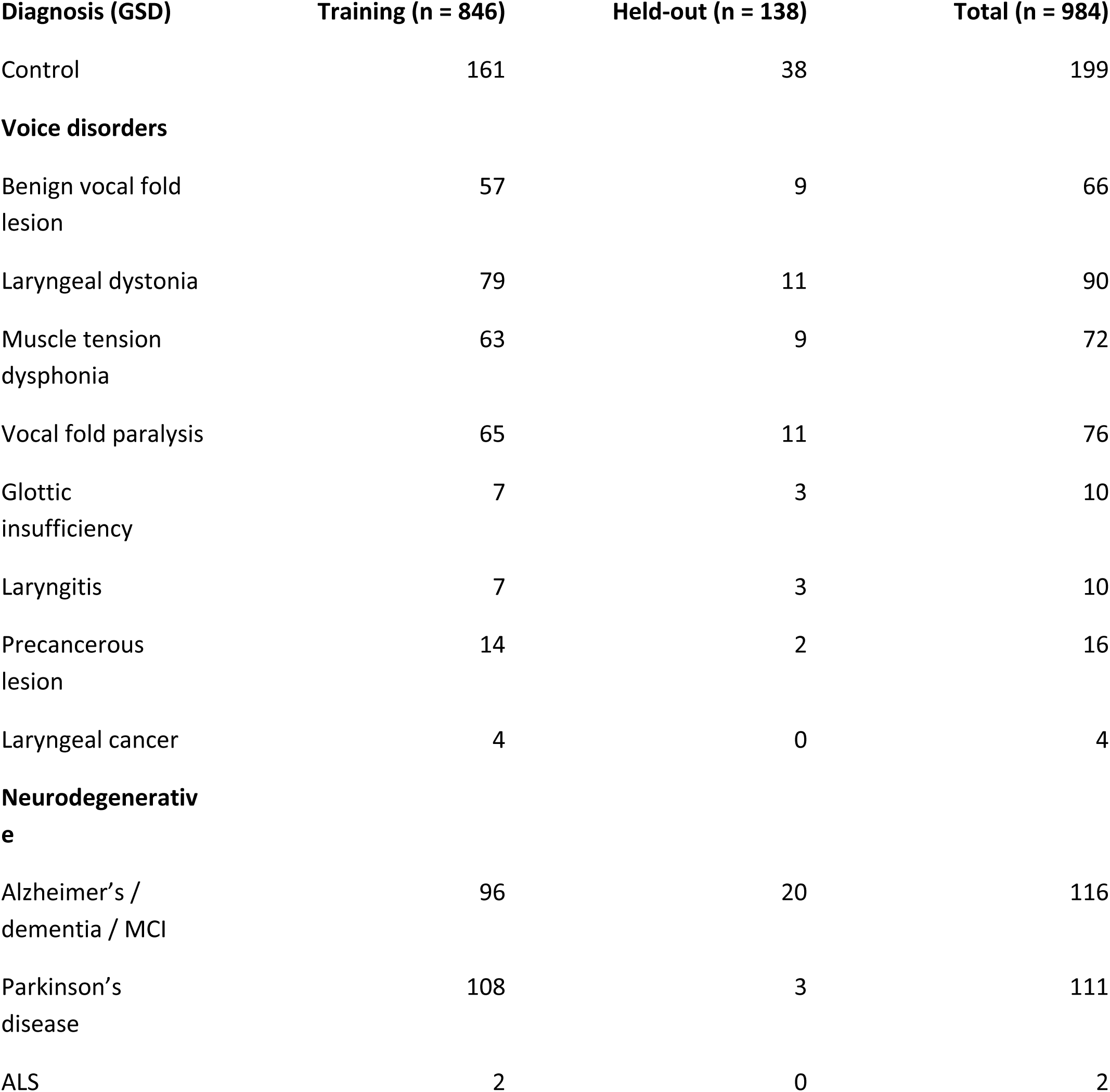
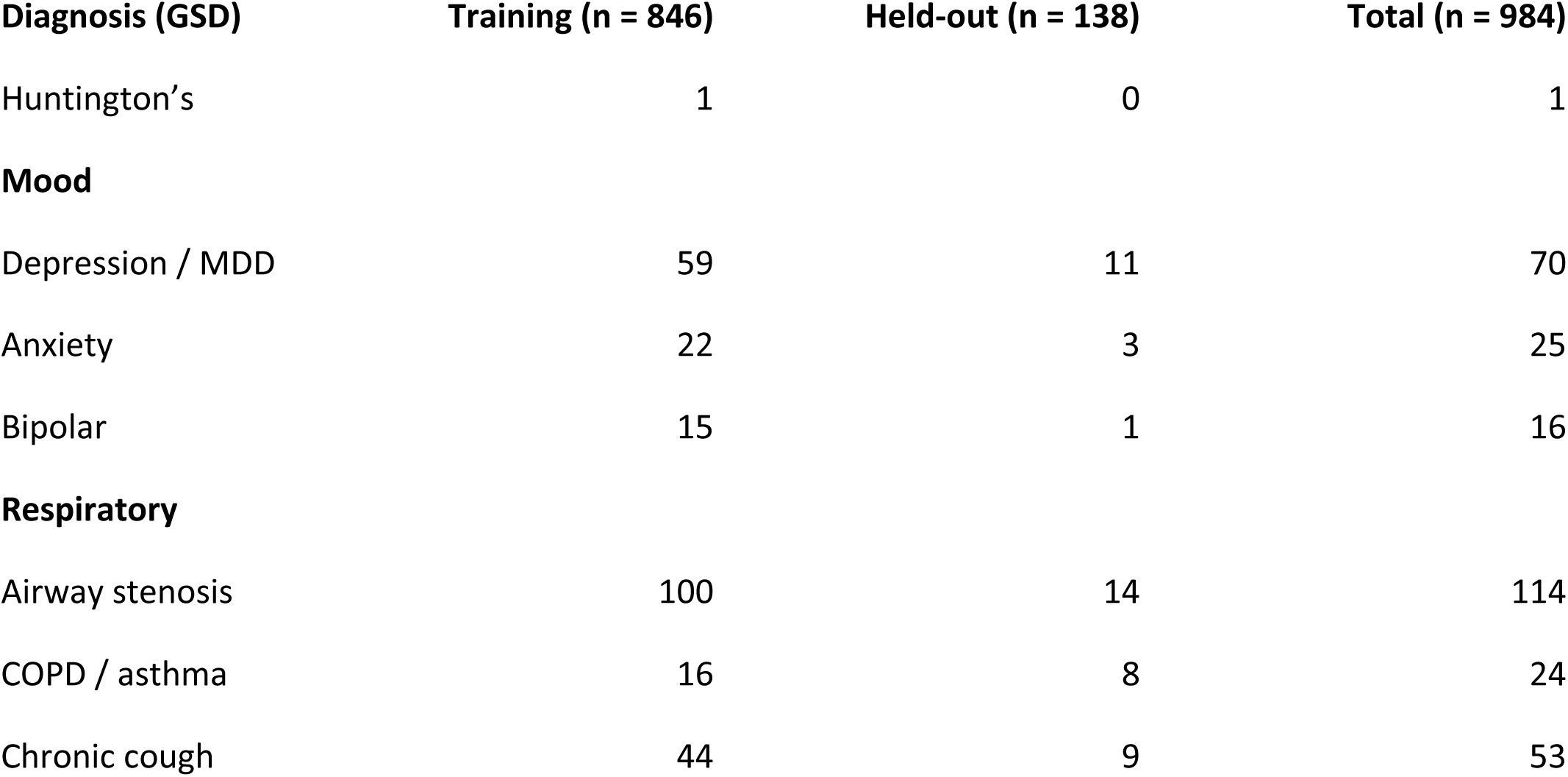
Per-diagnosis cohort sizes. Participant counts per clinician-validated gold-standard diagnosis (GSD), broken down by cohort partition. A participant may carry more than one GSD; counts therefore sum to more than the cohort size. Self-report counts may differ from the clinician-validated GSD numbers reported here. Note: disease-category counts cited in the main text (e.g., voice n = 333, mood n = 95) are unique participants per category (any-GSD-positive); the per-GSD rows below are per-diagnosis participant counts and may sum to more than the category total because some participants carry multiple GSDs within the same category.

**Figure S1.**
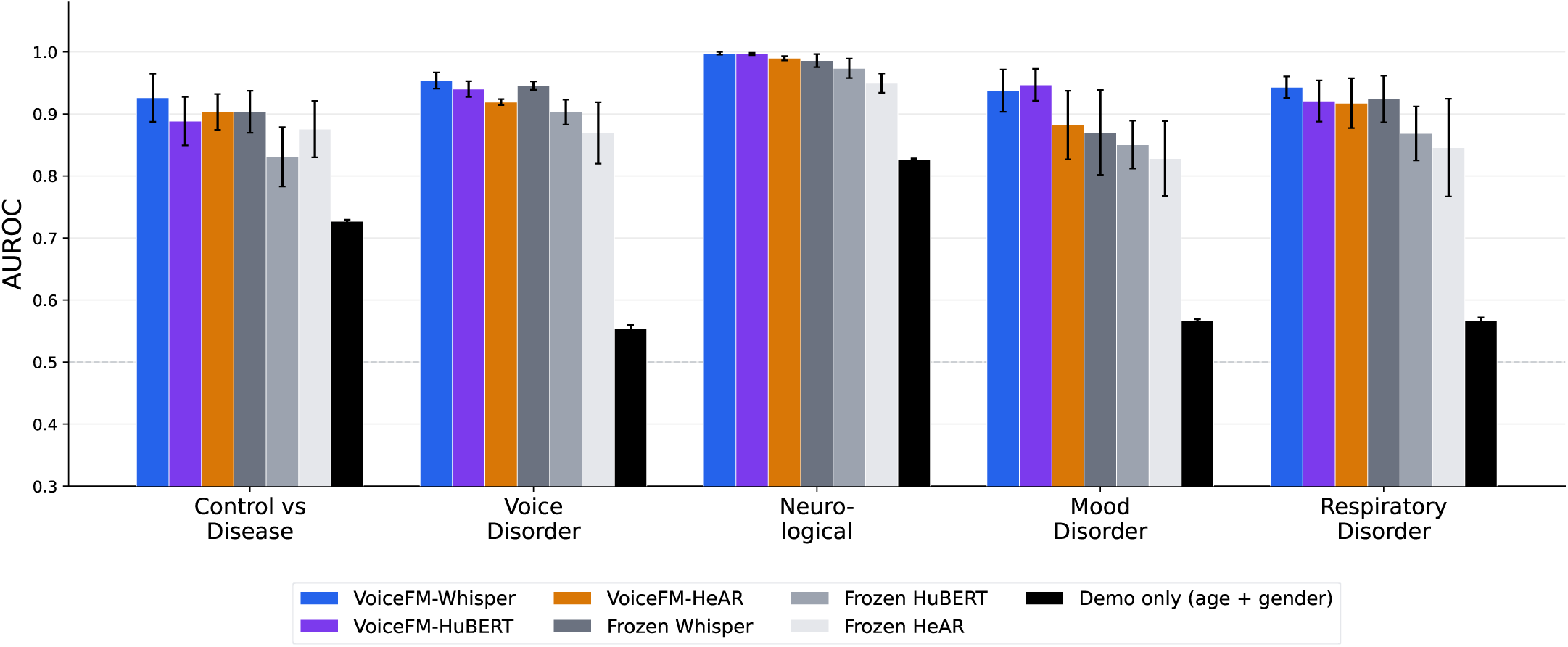
Full model comparison and demographic baseline. GSD category AUROC for six audio-embedding models — VoiceFM-Whisper, VoiceFM-HuBERT, VoiceFM-HeAR, Frozen Whisper, Frozen HuBERT, Frozen HeAR — and a demographics-only baseline (black bars: logistic regression on age + sex alone, no audio). VoiceFM-HeAR uses a frozen Google HeAR encoder with only the projection layer trainable. The demographic baseline is shown to rule out the possibility that AUROC is driven by demographic confounding rather than acoustic disease information. Error bars: standard deviation across 5 seeds (42–46).

**Figure S2.**
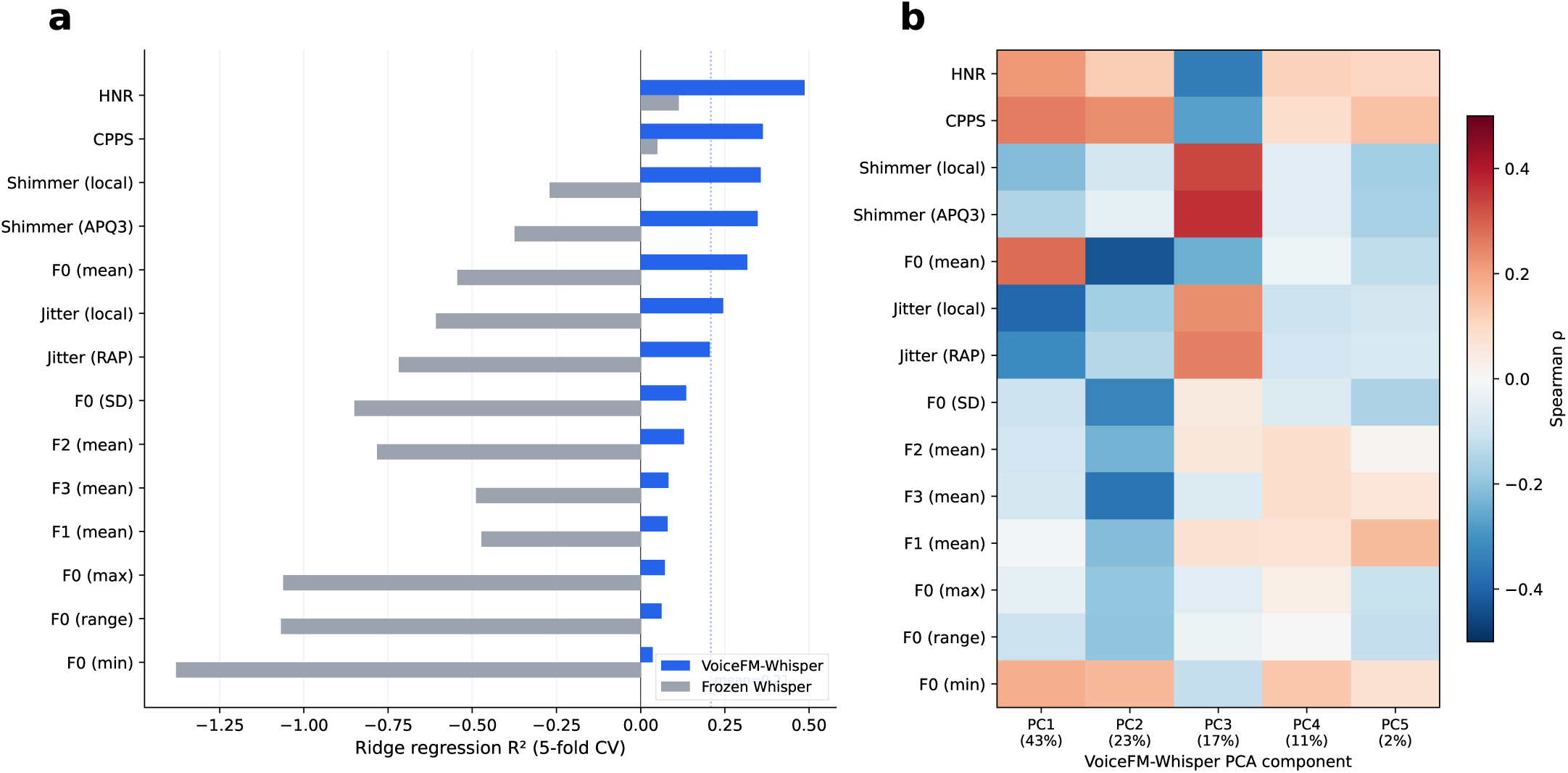
Acoustic grounding (interpretability) a, Ridge regression R^2^ (5-fold CV, scaler fit inside fold) for 14 traditional acoustic features decoded from VoiceFM-Whisper (blue) vs Frozen Whisper (gray) embeddings. b, Spearman rank correlation between each acoustic feature and the top PCA components of VoiceFM-Whisper embeddings. PC1 separates healthy-voice markers (CPPS, HNR) from pathological-voice markers (jitter, shimmer).

**Figure S3.**
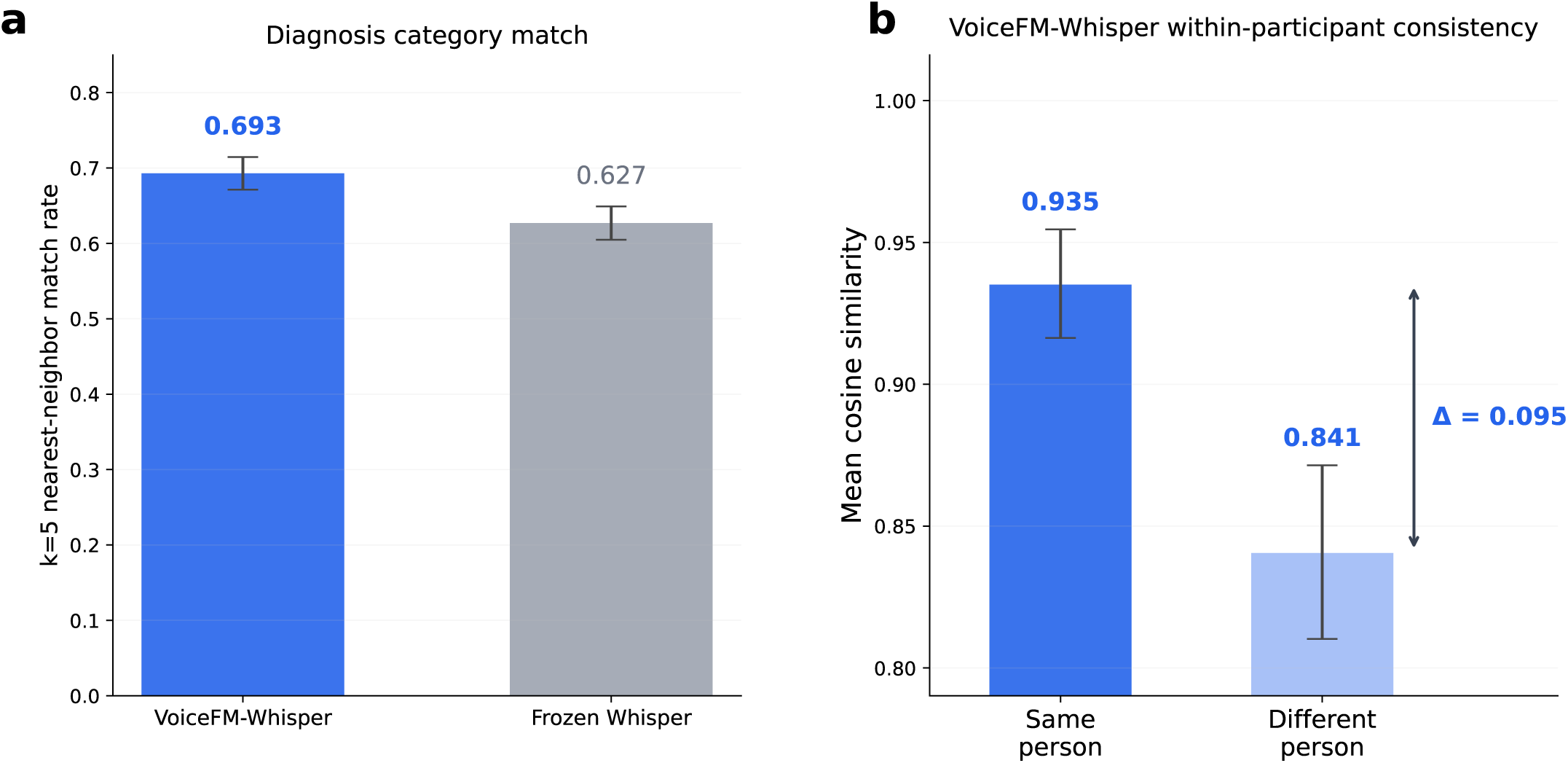
Embedding structure. a, Nearest-neighbor retrieval (k=5, cosine distance) on the 846 training participants: fraction of NNs sharing the query participant’s GSD category, for VoiceFM-Whisper vs Frozen Whisper. The chance baseline (random pair sharing a category) is ≍0.24. Welch’s t-test on the per-participant match rates is significant (p = 3.0×10^−5^). Error bars: 95% CI of the mean. b, Within-participant consistency for VoiceFM-Whisper: mean cosine similarity between embeddings of different recording types from the same participant (intra-person) vs different participants (inter-person). Bars show per-participant means; error bars show ±1 SD across the 845 training participants who have ≥2 recordings.

## References

[1] Pizzimenti M, Kalia A, Toghranegar JA, et al. Consensus-based definitions for vocal biomarkers: the International VOCAL Initiative. medRxiv. 2025. doi:10.1101/2025.10.23.25338518

[2] Fagherazzi G, et al. Voice for Health: The Use of Vocal Biomarkers from Research to Clinical Practice. Digit Biomark. 2021;5(1):78–88. doi:10.1159/000515346

[3] Harel BT, et al. Acoustic characteristics of Parkinsonian speech: a potential biomarker of early disease progression and treatment. J Neurolinguistics. 2004;17(6):439–453. doi:10.1016/j.jneuroling.2004.06.001

[4] Cummins N, et al. A review of depression and suicide risk assessment using speech analysis. Speech Commun. 2015;71:10–49. doi:10.1016/j.specom.2015.03.004

[5] Hillenbrand J, Houde RA. Acoustic correlates of breathy vocal quality: Dysphonic voices and continuous speech. J Speech Hear Res. 1996;39(2):311–321. doi:10.1044/jshr.3902.311

[6] Tracy JM, et al. Investigating voice as a biomarker: Deep phenotyping methods for early detection of Parkinson’s disease. J Biomed Inform. 2020;104:103362. doi:10.1016/j.jbi.2019.103362

[7] Fraser KC, et al. Linguistic features identify Alzheimer’s disease in narrative speech. J Alzheimers Dis. 2016;49(2):407–422. doi:10.3233/JAD-150520

[8] Bagad P, et al. Cough against COVID: Evidence of COVID-19 signature in cough sounds. arXiv preprint arXiv:2009.08790. 2020.

[9] Al-Nasheri A, Muhammad G, Alsulaiman M, et al. Voice pathology detection and classification using auto-correlation and entropy features in different frequency regions. IEEE Access. 2018;6:6961–6974. doi:10.1109/ACCESS.2017.2696056

[10] Bensoussan Y, Elemento O, Rameau A. Voice As an AI Biomarker of Health — Introducing Audiomics. JAMA Otolaryngol Head Neck Surg. 2024;150(4):283–284. doi:10.1001/jamaoto.2023.4807

[11] Robin J, et al. Evaluation of speech-based digital biomarkers: Review and recommendations. Digit Biomark. 2020;4(3):99–108. doi:10.1159/000510820

[12] Azizi S, et al. Big self-supervised models advance medical image classification. In: Proc. IEEE/CVF ICCV. 2021:3458–3468. doi:10.1109/ICCV48922.2021.00346

[13] Steinberg E, et al. Language models are an effective representation learning technique for electronic health record data. J Biomed Inform. 2021;113:103637. doi:10.1016/j.jbi.2020.103637

[14] Jumper J, et al. Highly accurate protein structure prediction with AlphaFold. Nature. 2021;596(7873):583–589. doi:10.1038/s41586-021-03819-2

[15] Hsu WN, et al. HuBERT: Self-supervised speech representation learning by masked prediction of hidden units. IEEE/ACM Trans Audio Speech Lang Process. 2021;29:3451–3460. doi:10.1109/TASLP.2021.3122291

[16] Baevski A, et al. wav2vec 2.0: A framework for self-supervised learning of speech representations. NeurIPS. 2020.

[17] Baur S, Nabulsi Z, Weng WH, et al. HeAR — Health Acoustic Representations. arXiv preprint arXiv:2403.02522. 2024. Available at: https://huggingface.co/google/hear-pytorch

[18] Bensoussan Y, Sigaras A, Rameau A, Elemento O, et al. Bridge2AI-Voice: An ethically-sourced, diverse voice dataset linked to health information (version 3.0.0). PhysioNet. 2025. doi:10.13026/k81f-qr68

[19] Radford A, et al. Learning transferable visual models from natural language supervision. In: Proc. ICML. PMLR 139; 2021:8748–8763.

[20] Huang X, Khetan A, Cvitkovic M, Karnin Z. TabTransformer: Tabular data modeling using contextual embeddings. arXiv preprint arXiv:2012.06678. 2020.

[21] van den Oord A, Li Y, Vinyals O. Representation learning with contrastive predictive coding. arXiv preprint arXiv:1807.03748. 2018.

[22] Radford A, et al. Robust speech recognition via large-scale weak supervision. In: Proc. ICML. PMLR 202; 2023:28492–28518.

[23] Mendes-Laureano J, Gómez-García JA, Guerrero-López A, Luque-Buzo E, Arias-Londoño JD, Grandas-Pérez FJ, Godino-Llorente JI. NeuroVoz: a Castillian Spanish corpus of parkinsonian speech. Sci Data. 2024;11:1367. doi:10.1038/s41597-024-04186-z

[24] Eyben F, Scherer KR, Schuller BW, et al. The Geneva Minimalistic Acoustic Parameter Set (GeMAPS) for voice research and affective computing. IEEE Trans Affect Comput. 2016;7(2):190–202. doi:10.1109/TAFFC.2015.2457417

[25] Eyben F, Wöllmer M, Schuller B. openSMILE — The Munich versatile and fast open-source audio feature extractor. In: Proceedings of the 18th ACM International Conference on Multimedia (MM ‘10). 2010:1459–1462. doi:10.1145/1873951.1874246

[26] Bot BM, et al. The mPower study, Parkinson disease mobile data collected using ResearchKit. Sci Data. 2016;3:160011. doi:10.1038/sdata.2016.11

[27] Wroge TJ, et al. Parkinson’s disease diagnosis using machine learning and voice. In: 2018 IEEE Signal Processing in Medicine and Biology Symposium (SPMB). 2018:1–7. doi:10.1109/SPMB.2018.8615607

[28] Rahmatallah Y, Kemp AS, Iyer A, Pillai L, Larson-Prior LJ, Virmani T, Prior F. Pre-trained convolutional neural networks identify Parkinson’s disease from spectrogram images of voice samples. Sci Rep. 2025;15:7337. doi:10.1038/s41598-025-92105-6

[29] Tougui I, Zakroum M, Karrakchou O, Ghogho M. Transformer-based transfer learning on self-reported voice recordings for Parkinson’s disease diagnosis. Sci Rep. 2024;14:30131. doi:10.1038/s41598-024-81824-x

[30] Williamson JR, Quatieri TF, Helfer BS, Perricone J, Ghosh SS, Ciccarelli G, Mehta DD. Segment-dependent dynamics in predicting Parkinson’s disease. Proc Interspeech 2015. 2015:518–522. doi:10.21437/Interspeech.2015-187

[31] Awan SN, Bensoussan Y, Watts S, Boyer M, Budinsky R, Bahr RH. Influence of recording instrumentation on measurements of voice in sentence contexts: use of smartphones and tablets. Front Digit Health. 2025;7:1610772. doi:10.3389/fdgth.2025.1610772

[32] Sharma N, et al. Coswara — A database of breathing, cough, and voice sounds for COVID-19 diagnosis. In: Proc. Interspeech 2020. 2020:4811–4815. doi:10.21437/Interspeech.2020-2768

[33] Pützer M, Barry WJ. Saarbrücken Voice Database [Internet]. Institute of Phonetics, Saarland University. Available from: https://stimmdb.coli.uni-saarland.de/

[34] Jaeger H, Trivedi D, Stadtschnitzer M. Mobile device voice recordings at King’s College London (MDVR-KCL) from both early and advanced Parkinson’s disease patients and healthy controls. Zenodo. 2019. doi:10.5281/zenodo.2867215

[35] Kornblith S, et al. Similarity of neural network representations revisited. In: Proc. ICML. PMLR 97; 2019:3519–3529.

[36] Collins GS, Moons KGM, Dhiman P, et al. TRIPOD+AI statement: updated guidance for reporting clinical prediction models that use regression or machine learning methods. BMJ. 2024;385:e078378. doi:10.1136/bmj-2023-078378

